# ‘EVOLVE-HBV’: A retrospective cross-sectional study to quantify and characterise HBV infection, exposure, immunity and susceptibility in a rural population in KwaZulu-Natal, South Africa

**DOI:** 10.64898/2026.03.17.26347919

**Authors:** Motswedi Anderson, Lusanda Mazibuko, Gloria Sukali, Tongai G Maponga, Marion Delphin, Elizabeth Waddilove, Janine Upton, VG Naidoo, Stephen Olivier, Gregory Ording-Jespersen, Dickman Gareta, Emily Martyn, Resign Gunda, Kobus Herbst, Willem A Hanekom, Nokukhanya Msomi, Lulama Mthethwa, Malcolm Ellapen, Theresa Smit, Thumbi Ndung’u, Emily B Wong, Mark J Siedner, Thandeka Khoza, Kathy Baisley, Collins Iwuji, Philippa C Matthews

**Affiliations:** Africa Health Research Institute, KwaZulu Natal, South Africa; Botswana Harvard Health Partnership Institute, Gaborone, Botswana; The Francis Crick Institute, 1 Midland Road, London NW1 1AT London, UK; Division of Infection and Immunity, University College London, London, WC1E 6BT, UK; Division of Medical Virology, Stellenbosch University, Faculty of Medicine and Health Sciences, Tygerberg, Cape Town, South Africa; National Health Laboratory Service, Tygerberg Business Unit, Tygerberg, Cape Town, South Africa; Department of Gastroenterology and Hepatology, Inkosi Albert Luthuli Central Hospital, Durban, South Africa; DSTI-SAMRC South African Population Research Infrastructure Network (SAPRIN), Durban, South Africa; Discipline of Virology, School of Medicine, University of KwaZulu–Natal, South Africa; National Health Laboratory Service, Inkosi Albert Luthuli Central Hospital, Durban, South Africa; Division of Infectious Diseases, Department of Medicine, University of Alabama at Birmingham, Birmingham, AL, USA; Division of Infectious Diseases, Massachusetts General Hospital, Boston, MA, USA; London School of Hygiene and Tropical Medicine, London, UK; Department of Global Health & Infection, Brighton and Sussex Medical School, University of Sussex, Brighton, BN1 9PX, UK; Department of Infectious Diseases, University College London Hospital, Euston Road, London NW1 2BU, UK

**Keywords:** HBV, hepatitis B, South Africa, screening, diagnosis, infection, vaccine, exposure, prevalence, epidemiology

## Abstract

**Introduction:** International goals aim to eliminate Hepatitis B Virus (HBV) as a public health threat by 2030, but data representing African populations remain limited. We therefore investigated the population prevalence of HBV and treatment eligibility in a rural South African setting.

**Methods:** We tested archived plasma samples from 2200 participants in a population-based study in KwaZuku Natal for HBV surface antigen (HBsAg), HBV core antibody (anti-HBc), and HBV surface antibody (anti-HBs). For samples testing HBsAg-positive, we quantified alanine transferase (ALT) and HBV DNA viral load. We evaluated demographic and clinical correlates of HBV biomarkers, explored the geographical distribution of HBsAg, and assessed HBV treatment eligibility.

**Results:** Weighted HBV infection prevalence was 10.4% (95% CI: 9.0%-12.1%). Evidence of HBV exposure and clearance was found in 34.9% (95% CI: 32.4 - 37.5). Overall prevalence of vaccine-mediated HBV immunity was 8.9% (95% CI: 7.5%-10.4%) but for the sub-group born between 2000-2005 (after the HBV vaccine was implemented) this increased to 20.2% (95% CI 15.8-25.4). Infection prevalence was highest in the South of the region. Over 60% of individuals testing HBsAg-positive met treatment eligibility criteria.

**Conclusion:** Prevalence of HBV infection and exposure in this setting is high, while vaccine-mediated immunity is low. These data highlight a pressing need for scale-up of interventions to support progress towards global elimination targets.

**Funding:** The Francis Crick Institute (ref. CC2223), the Africa Oxford Initiative (Research Development Award) and Wellcome Strategic Core Award (227167/A/23/z).

**Ethics:** University of KZN (UKZN) ref. 00004495/2022; University College London ref. 23221/001.

## 1.0 BACKGROUND

An estimated 254 million people worldwide are living with chronic hepatitis B virus (HBV) infection, of whom 65 million are in the World Health Organization (WHO) African region^1^. High profile international elimination targets for viral hepatitis include an increase in diagnosis and treatment to deliver reductions in morbidity and mortality by 2030^2^. By 2022, progress was still off track, with only 13% of people living with HBV (PLWHB) having been diagnosed (vs. 90% goal for 2030) and 3% receiving therapy (vs. 80% goal for 2030)^2^. WHO recommends that in areas where HBV prevalence is high in the general population (≥5%), adults should be routinely screened^3^. In South Africa (SA), estimates for the number of PLWHB vary, from 1.9 million^4^ to 3.5 million^5^, making it a middle to high prevalence area.

Robust, safe and affordable interventions are available for HBV prevention and treatment. Vaccine is recommended as a birth dose (BD) followed by routine infant immunisation from the age of 6 weeks as part of the WHO Expanded Programme for Immunisation^6^. This intervention has reduced HBV prevalence in children under 5 years old to <1% worldwide by 2022^7^, but vaccine coverage varies by region. In SA, HBV immunization programmes started in 1995 and vaccination coverage was reported to be between 67-86% between 2000 and 2019^4^.

Chronic HBV infection can be suppressed using nucleos(t)ide analogue (NA) agents, which are also used as antiretroviral therapy (ART) against human immunodeficiency virus (HIV). Tenofovir disoproxil fumarate (TDF) is the most common first-line agent for HBV^8^ and part of the recommended first-line ART regimen for HIV in most of sub-Saharan Africa. Until recently, treatment was only recommended in a minority of PLWHB, deemed at the highest risk of developing liver disease^8^. In many settings, risk stratification is difficult and therefore therapy was not consistently provided. However, new WHO guidelines published in 2024 recommend more inclusive and simplified treatment eligibility criteria, such that a higher proportion of PLWHB qualify for treatment^3^. This expansion of treatment will be an important tool for advancing towards elimination goals, targeting both the individual consequences of infection, and the population reservoir to reduce the risk of transmission.

Assessment of the prevalence of HBV infection, burden of associated liver disease, and treatment eligibility now frame viral hepatitis within a ‘syndemic’ context, characterised by the interaction of HBV with other infections and non-communicable diseases (NCDs) and recognising overlapping psychosocial factors^9^. In SA, 7.7 million people are living with HIV (PLWH)^10^, which can influence the prevalence of HBV markers and HBV viraemia, liver disease progression, and access to timely treatment. NCDs in PLWHB may also influence liver disease outcomes^11^; in particular, there is growing concern about metabolic liver disease which intersects with cardiometabolic risk factors (obesity, dyslipidemia, hypertension and diabetes). These conditions can increase the risk of liver fibrosis, cirrhosis, and hepatocellular carcinoma (HCC)^12^.

HBV has been neglected as a threat in the WHO African region^13,14^. To provide an evidence-base for allocation of resources and interventions, and to benchmark progress towards elimination targets, it is crucial to establish reliable estimates of HBV infection and immunity, and to investigate the burden and characteristics of infection and associated liver disease. We therefore set out to study a SA population located in rural Kwazulu-Natal (KZN), to (i) quantify the prevalence of HBV infection, exposure, vaccine-mediated immunity and susceptibility; (ii) determine associations between HBV serostatus and other clinical and demographic characteristics; (iii) assess HBV treatment eligibility based on recently revised WHO criteria.

## 2.0 METHODS

### 2.1 Study population

Our study was nested within the ‘Vukuzazi’ programme, a partnership between the Africa Health Research Institute (AHRI) and the uMkhanyakude community in KZN (SA)^15^ (**Figure 1A, Supplementary methods 1**). This programme included a community-wide health screening for HIV, tuberculosis, hypertension and diabetes, along with referral into primary care for those newly diagnosed with any of these conditions. In SA, HIV diagnosis and treatment is funded by state services. However, while HBV testing and treatment may be offered *ad hoc*, there are no unified, systematic pathways to access HBV testing and treatment, and access to care may impose out-of-pocket costs on individuals. To develop translational research focusing on HBV, we established a new project, ‘EVOLVE-HBV’ (Evaluation of Vukuzazi LiVEr disease - Hepatitis B) (**Figure 1B**).

**Figure 1:**
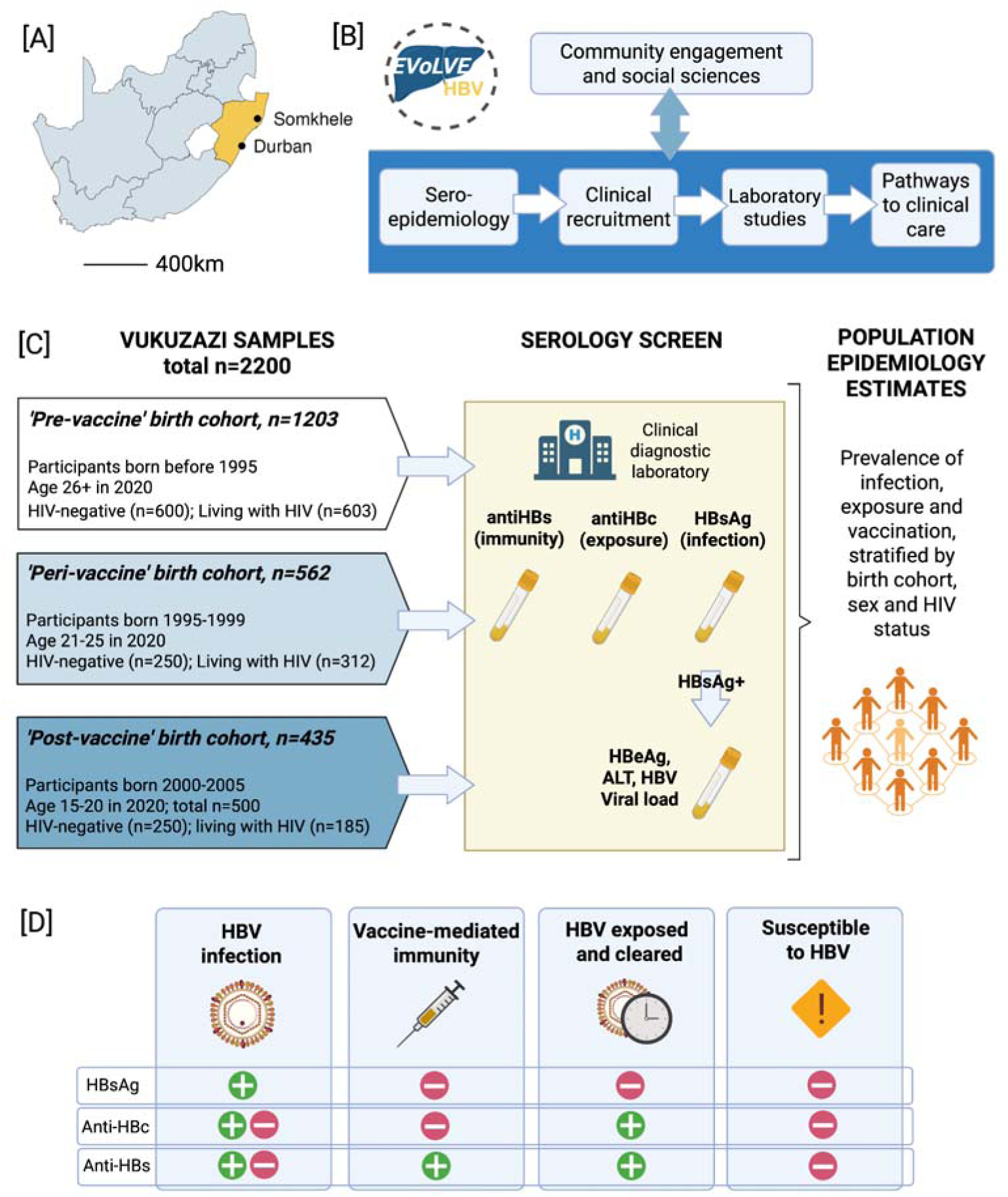
Schematic strategy of the EVOLVE-HBV programme exploring HBV seroepidemiology. [A] Map of South Africa highlighting KwaZulu-Natal province (gold) and AHRI site locations; [B] Flow diagram of EVOLVE-HBV study; [C] Selection of samples for screening HBV biomarkers; [D] interpretation of serology markers to determine HBV infection, vaccine-mediated immunity, HBV exposure and clearance, and susceptibility to HBV. Green ‘+’ means positive result, Red ‘-’ means negative result. AHRI: African Health Research Institute; EVOLVE-HBV: Evaluation of Vukuzazi LiVEr disease - Hepatitis B; HBV: Hepatitis B Virus; anti-HBc: HBV core antibodies; HBsAg: HBV surface antigen; anti-HBs: HBsAg antibodies; HIV: Human Immunodeficiency Virus. Figure panel A created in R studio, and panels B-D created in BioRender.com with a licence to publish.

### 2.2 Ethics and approvals

EVOLVE-HBV was approved by the University of KZN (UKZN) Biomedical Research Ethics Committee (ref. 00004495/2022) and University College London (UCL) ethics committee in the United Kingdom (UK) (ref. 23221/001). The study was also approved by the AHRI Community Advisory Board, and the Vukuzazi Scientific Steering Committee.

### 2.3 Sample selection

We selected a subset of 2200 plasma samples previously collected by the Vukuzazi programme to screen for HBV markers, stratified by age group (birth cohort), sex, and HIV status (**Figure 1C**). The samples had been collected between May 2018 and March 2020 from participants who were aged ≥15 years at the time of enrolment, and stored at -80°C. Our sample selection was designed to represent population groups born in different ‘epochs’ according to HBV vaccination schedules:

**(i) Pre-vaccine roll-out** (born before 1995)
**(ii) Peri-vaccine roll-out** (born 1995-1999)
**(iii) Post vaccine-roll out** (born 2000-2005).

Within these groups, samples were selected at random, subject to availability of sufficient sample volume.

### 2.4 Laboratory testing and classification of HBV serostatus

Plasma samples were screened for HBV biomarkers of infection, exposure and immunity. We defined HBV serological groups as follows **(Figure 1D):**

**(i) HBV infection** (defined as HBsAg positive, irrespective of other markers);
**(ii) HBV exposure with clearance** (defined as HBsAg-negative with anti-HBc positive, in the presence or absence of anti-HBs);
**(iii) Vaccine-mediated immunity** (defined as anti-HBs positive in the absence of any other seromarkers);
**(iv) Susceptible to HBV infection** (negative for all HBV markers).

Primary assay details are presented in **Suppl Table 1.** Samples that tested HBsAg-positive were further tested for HBV e-antigen (HBeAg), alanine aminotransferase (ALT) and HBV DNA viral load (VL) levels. 20 selected samples reported as HBsAg-positive were also retested on two alternative platforms (**Suppl methods 2 and Suppl Table 2**).

### 2.5 Application of 2024 WHO HBV treatment guidelines

We applied the 2024 WHO HBV guidelines to determine which individuals would meet eligibility criteria for HBV treatment^3^ (criteria in **Suppl Table 3**). Based on the data available for this population, we first narrowed those eligible to a positive HBsAg test based on our primary diagnostic assay (Abbott; HBsAg S/CO>1.0). Among the population not currently treated, we considered those potentially eligible based on HBV DNA >2000 IU/mL and ALT >ULN, or the presence of a comorbidity or coinfection (diabetes, HIV), irrespective of HBV DNA or ALT quantification. We repeated assessment for participants meeting a more stringent diagnostic threshold of HBsAg S/CO >10 which is an alternative cut-off that has been suggested to improve specificity^16,17^.

### 2.6 Statistical analysis

Full details of the statistical methods are in **Suppl Methods 3**. In brief, to generate prevalence estimates, we employed sampling weights to account for the unequal probability of selection from the Vukuzazi sample bank. Weights were calculated as the inverse probability of selection within each age, sex and HIV status stratum. In addition, we applied non-response weights to adjust for differential rates of participation in the Vukuzazi programme; weights were calculated as the inverse probability of participation by sex and age group (15-24, 25-44, 45-64 and ≥65 years)^18^.

To describe the characteristics of our study sample, we used frequencies and proportions for categorical variables, and medians and interquartile ranges (IQR), or means and standard deviations (SD), for continuous variables. We estimated the weighted overall prevalence of our four serological categories (HBV infection / exposure with clearance / vaccine-mediated immunity / susceptibility). This weighting allowed us to produce estimates generalisable to the whole surveillance population, and to allow valid inference. Unweighted estimates would not provide this information, because the age, sex and HIV status distribution in the sample, by design, does not reflect that of the general population. We produced continuous surface maps of weighted HBV infection prevalence, based on location of residence within the surveillance area at the time of sample collection. We used a standard Gaussian kernel interpolation method with a search radius of 2km, to identify potential geographic disparities in HBV infection.

We fitted weighted logistic regression models to estimate odds ratios (OR) and 95% confidence intervals (CI) for factors associated with each HBV serological category. All models were adjusted for the stratification variables (birth cohort, sex and HIV status) and the following additional covariates: education, socioeconomic status (SES), alcohol intake, smoking status, BMI category, HTN and diabetes. Definition and thresholds are in **Suppl methods 4**.

In addition to estimating HBsAg prevalence based on the manufacturer’s instructions using HBsAg S/CO>1, we also repeated adjusted regression using more stringent HBsAg S/CO cut-offs (>2, >5, >10, >100), reporting sensitivity analysis using S/CO >10.

### 2.7 Software and availability of data and code

Analyses were conducted in Stata version 18 and R version 4.1.3. Data for this analysis are archived in the AHRI data repository: {link to data will be provided on acceptance}. The R code used for this analysis is available here: {link to code will be provided on acceptance}.

## 3.0 RESULTS

### 3.1 Sociodemographic and clinical characteristics

Reflecting our sampling strategy, our study cohort (n=2200) was 55.3% female and 50.0% were PLWH (**Table 1**). At the time of sample collection, the median age was 27 years (IQR 20.0 - 45.0) with 54.7% born before 1995, 25.5% between 1995-1999, and 19.8% between 2000-2005. Among PLWH, only 72.5% were receiving ART, but among those on ART, 96.1% (767/798) had HIV viraemic suppression (corresponding to 69.7% suppression among all PLWH). The most commonly used ART combination was tenofovir/emtricitabine (TDF/FTC). Diabetes, hypertension, being overweight and obesity were documented in 6.2%, 16.6%, 22.4%, and 21.8% respectively.

**Table 1:** Cohort description: sample characteristics of 2200 individuals included in the EVOLVE HBV serosurvey from the Vukuzazi programme in rural kwaZulu-Natal, South Africa. This table presents raw / unweighted data.

| Characteristic |  | Number (%) |
| --- | --- | --- |
| Birth Epoch | Born before 1995 | 1203 (54.7%) |
|  | Born 1995-1999 | 562 (25.5%) |
|  | Born 2000-2005 | 435 (19.8%) |
| Sex | Male | 984 (44.7%) |
|  | Female | 1216 (55.3%) |
| Education level | No formal education | 120 (5.5%) |
|  | Primary | 302 (13.7%) |
|  | Secondary | 1407 (63.9%) |
|  | Tertiary | 370 (16.8%) |
|  | Missing | 1 (<0.1%) |
| SES | Q1 (Lowest) | 240 (10.9%) |
|  | Q2 | 601 (27.3%) |
|  | Q3 | 517 (23.5%) |
|  | Q4 | 374 (17.0%) |
|  | Q5 (Highest) | 395 (18.0%) |
|  | Missing | 73 (3.3%) |
| Alcohol intake | Never drinker | 1820 (82.7%) |
|  | No drinking in last 12 months | 48 (2.2%) |
|  | Drinking in past 12 months | 332 (15.1%) |
| Smoking status | Never smoker | 1959 (89.0%) |
|  | Current smoker | 221 (10.0%) |
|  | Ex-smoker | 20 (0.9%) |
| HIV status | Negative | 1100 (50.0%) |
|  | Positive | 1100 (50.0%) |
| On ART (if living with HIV) | No | 302 (27.5%) |
|  | Yes | 798 (72.5%) |
| HIV viral suppression status | Not suppressed | 333 (30.3%) |
|  | Suppressed | 767 (69.7%) |
| BMI Category | Underweight | 139 (6.3) |
|  | Normal | 1076 (48.9%) |
|  | Overweight | 493 (22.4%) |
|  | Obese | 479 (21.8%) |
|  | Missing | 13 (0.6%) |
| Hypertension | No | 1830 (83.2%) |
|  | Yes | 365 (16.6%) |
|  | Missing | 5 (0.2%) |
| Diabetes | No | 2063 (93.8%) |
|  | Yes | 136 (6.2%) |
|  | Missing | 1 (<0.1%) |

ART: antiretroviral therapy; BMI: Body Mass Index; HIV: human immunodeficiency virus; SES: Socio Economic Status; HTN: hypertension.

### 3.2 Prevalence and combinations of HBV sero-markers

Based on our primary assay read-out of HBsAg S/CO >1, the overall weighted prevalence of HBV infection was 10.4% (95% CI: 9.0 - 12.1). The prevalence of HBV exposure and clearance was 34.9% (95% CI: 32.4 - 37.5), and vaccine-mediated immunity (anti-HBs >10 mIU/mL) 8.9% (95% CI: 7.5 - 10.4). Accordingly, nearly half of our study population was susceptible to HBV infection, weighted prevalence 45.8% (95% CI 43.2-48.4) (**Table 2**). We noted diverse combinations of serological markers: for example, among 240 samples testing HBsAg-positive, only 129 were also positive for anti-HBc, while 36 were positive for both HBsAg and anti-HBs (**Figure 2**).

**Figure 2:**
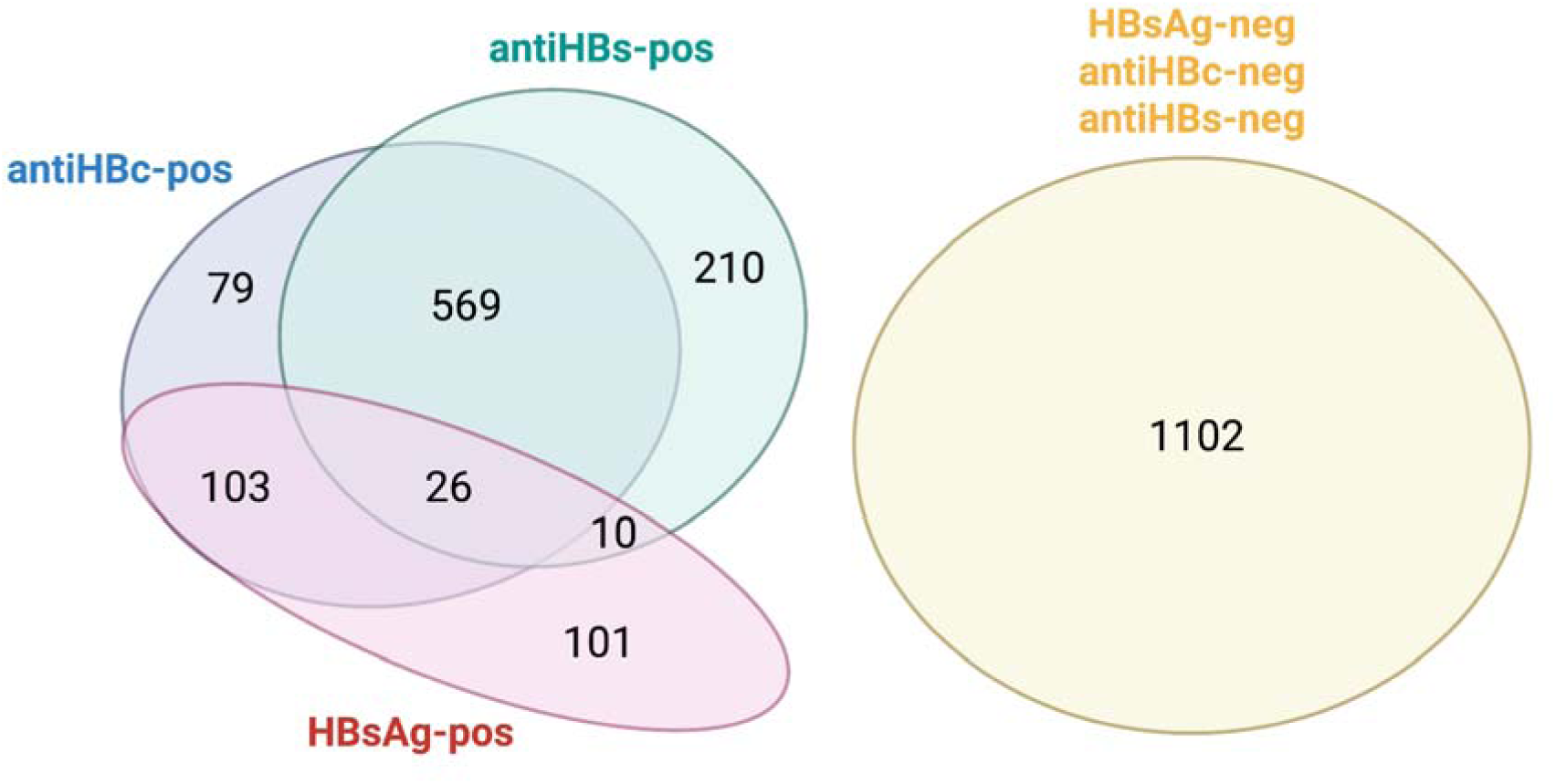
Summary of data from a serosurvey in the Kwa-Zulu Natal area of South Africa, showing the number of samples testing positive for HBV biomarkers, occurring individually and in combination. AntiHBc: HBV core antibodies; HBsAg: HBV surface antigen; antiHBs: HBV surface antibodies. Data shown for primary testing at National Health Laboratory Service laboratories, Albert Luthuli Hospital, Durban, with HBsAg defined by Abbott platform (positive defined as HBsAg S/CO ratio >1). Data shown for 2200 samples tested, with numbers in each category indicated.

**Table 2.** Weighted prevalence of HBV sero-categories stratified by sex, HIV status, and HBV birth epoch in a rural population in KwaZulu Natal, South Africa. Results based on primary testing at Albert Luthuli hospital clinical diagnostic laboratory with HBsAg S/CO>1.

| Variable | HBV infection | *p-value*<sup>1</sup> | HBV exposure & clearance | *p-value*<sup>1</sup> | Vaccine-mediated immunity to HBV | *p-value*<sup>1</sup> | HBV Susceptible | *p-value*<sup>1</sup> |
| --- | --- | --- | --- | --- | --- | --- | --- | --- |
|  | *Weighted prevalence (95 % CI)* |  | *Weighted prevalence (95 % CI)* |  | *Weighted prevalence (95 % CI)* |  | *Weighted prevalence (95 % CI)* |  |
| **Whole cohort** | 10.4<br>(9.0- 12.1) |  | 34.9<br>(32.4- 37.5) |  | 8.9<br>(7.5- 10.4) |  | 45.8<br>(43.2- 48.4) |  |
| **Sex** |  | 0.344 |  | 0.001 |  | 0.681 |  | 0.005 |
| Female | 9.8<br>(7.9- 12.2) |  | 38.2<br>(34.6- 41.9) |  | 9.1<br>(7.3- 11.3) |  | 42.8<br>(39.3- 46.4) |  |
| Male | 11.3<br>(9.3- 13.6) |  | 30.1<br>(27.0- 33.4) |  | 8.5<br>(6.7- 10.9) |  | 50.1<br>(46.5- 53.6) |  |
| **HBV vaccine age epoch** |  | 0.007 |  | <0.001 |  | <0.001 |  | <0.001 |
| Born < 1995 | 11.8<br>(10.0- 14.0) |  | 47.7<br>(44.4- 51.0) |  | 5.5<br>(4.1- 7.2) |  | 35.0<br>(31.9- 38.2) |  |
| Born 1995-1999 | 7.8<br>(5.4- 11.2) |  | 7.0<br>(5.0- 9.8) |  | 13.0<br>(9.8- 17.0) |  | 72.2<br>(67.3- 76.6) |  |
| Born 2000-2005 | 6.6<br>(4.3- 9.9) |  | 1.8<br>(0.8- 4.3) |  | 20.2<br>(15.8- 25.4) |  | 71.4<br>(65.8- 76.4) |  |
| **HIV status** |  | 0.027 |  | <0.001 |  | 0.022 |  | <0.001 |
| Negative | 9.2<br>(7.5- 11.3) |  | 29.6<br>(26.5- 32.9) |  | 10.0<br>(8.2- 12.1) |  | 51.1<br>(47.8- 54.4) |  |
| Positive | 12.8<br>(10.4- 15.7) |  | 45.1<br>(41.0- 49.2) |  | 6.7<br>(5.0- 8.9) |  | 35.4<br>(31.6- 39.4) |  |

<sup>1</sup>P-value from adjusted Wald test.
HIV: human immunodeficiency virus; HBV: hepatitis B virus; CI: confidence interval; S/CO: sample/cut-off

### 3.3 Factors associated with HBV infection

Among 240 participants with HBsAg, 6.7% (95% CI: 4.0 - 10.8) tested positive for HBeAg. HBV infection was higher in PLWH compared to the HIV-negative population on crude estimates (12.8% vs 9.2%; p=0.027; **Table 2**), but statistical significance was lost after adjustment (aOR 1.36, 95% CI 0.95-1.96; **Figure 3; Suppl Table 4**).

**Figure 3.**
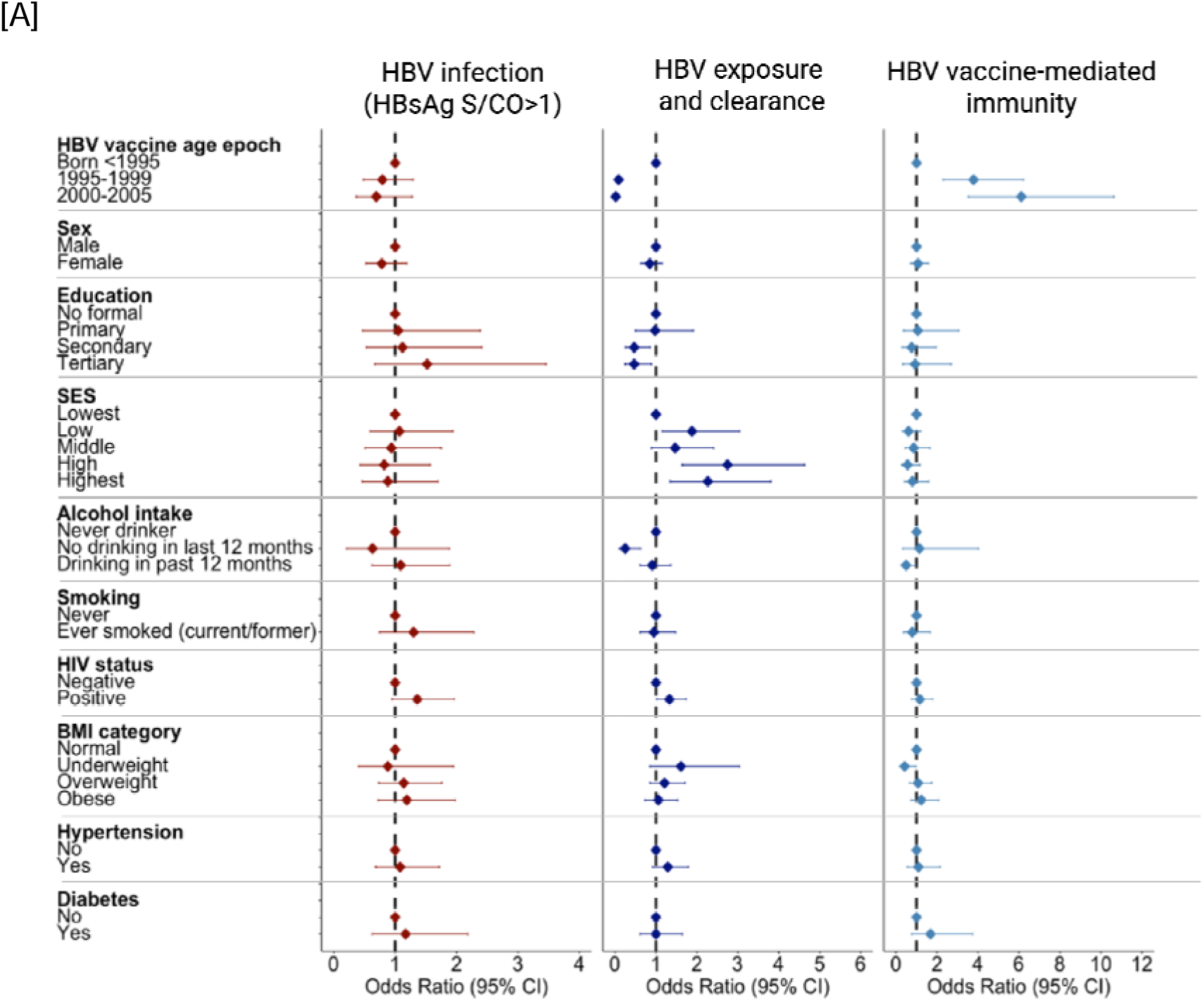
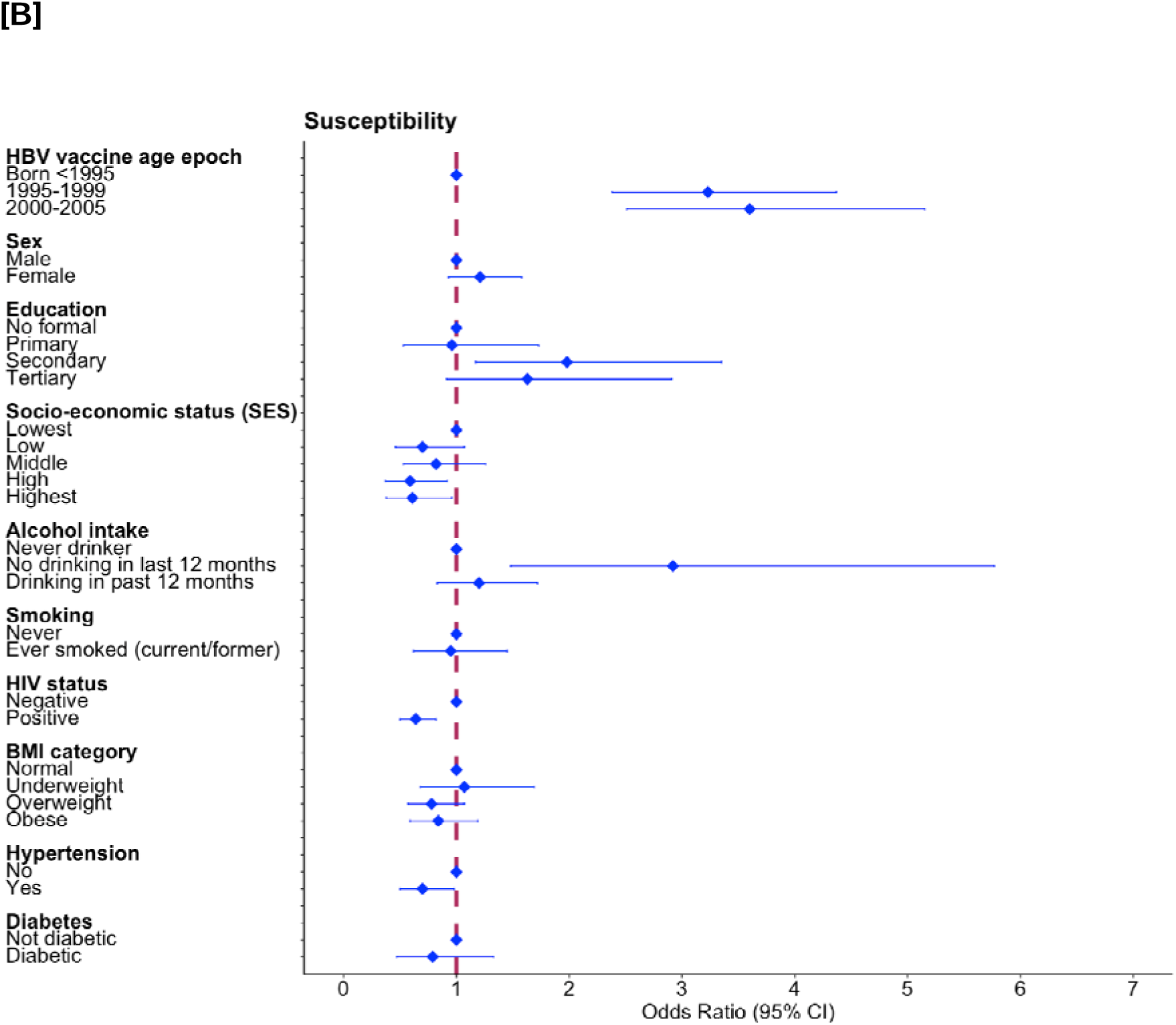
Forest plot of multivariable analysis of factors associated with HBV serostatus in 2200 adults sampled from the Vukuzazi cohort in KwaZulu Natal, South Africa. [A] HBV infection, exposure and immunity. [B] HBV susceptibility. HBV infection is defined based on laboratory report of HBsAg S/CO >1 and serological profiles are defined as shown in Figure 1D. HBV: Hepatitis B Virus; HIV: human immunodeficiency virus. BMI: Body mass index; S/CO: sample/cut-off. All numeric odds ratios and 95% CI are also presented in **Suppl Table 7**.

HBV infection prevalence did not differ by sex, either in crude estimates (9.8% in females vs 11.3% in males, p=0.34), or in adjusted models (female vs male aOR 0.78, 95% CI 0.52-1.19). Point estimates of HBV infection prevalence decreased across birth cohorts over time (from 11.8% in those born pre-1995, to 7.8% in those born 1995-1999, and 6.6% in those born from 2000-2005; p=0.007; **Table 2**), but, after adjustment, this statistical significance was lost (**Figure 3; Suppl Table 5**). When using a more stringent definition of infection (HBsAg S/CO>10), the adjusted odds of HBV infection remained significantly lower in younger cohorts (**Suppl Table 6-8**).

ALT was >ULN in 20.3% (14.9 - 26.9) with 17.9% (12.6 -24.7) being in those on ART versus 53.8% (29.6 -76.3) in those not on ART (**Table 3).** 53.8% (29.6 - 76.3) who tested HBeAg-positive had elevated ALT as compared to 17.9% (12.6 - 24.7) of those HBeAg-negative.

**Table 3:** Weighted estimates of laboratory parameters in individuals with HBsAg-positive status (n=240) stratified according to the presence of HIV co-infection and receipt of tenofovir-based antiretroviral therapy (ART)

| Variable | HBV mono-infection | HBV/HIV co-infection on ART | HBV/HIV co-infection not on ART |
| --- | --- | --- | --- |
| Number of individuals | 111 | 84 | 45 |
| HBV VL (IU/mL), mean (SD) | 358.9<br>(14.2) | 94.5<br>(34.1) | 14998.7<br>(418.0) |
| HBV VL category: <10 iU/mL | 48.9%<br>(38.2 - 59.7) | 69.8%<br>(57.8 - 81.8) | 29.9%<br>(9.7 - 50.1) |
| HBV VL category: 10-2,000 iU/mL | 36.4%<br>(25.7 - 47.0) | 25.3%<br>(13.9 - 36.7) | 29.7%<br>(12.4 - 47.0) |
| HBV VL category: 2,001-20,000 iU/mL | 10.0%<br>(3.4 - 16.6) | 1.3%<br>(0.0 - 3.8) | 19.2%<br>(4.2 - 34.1) |
| HBV VL category: >20,000 iU/mL | 4.7%<br>(0.7 - 8.7) | 3.6%<br>(0.0 - 8.1) | 21.2%<br>(5.4 - 37.1) |
| HBeAg status: Negative (95% CI) | 97.5%<br>(94.4 - 100.0) | 92.5%<br>(86.1 - 98.9) | 70.3%<br>(52.4 - 88.2) |
| HBeAg status: Positive (95% CI) | 2.5%<br>(0.0 - 5.6) | 7.5%<br>(1.1 - 13.9) | 29.7%<br>(11.8 - 47.6) |
| ALT (U/L), mean (SD) | 16.7 (8.6) | 24.5<br>(19.6) | 29.4<br>(17.6) |
| ALT category (WHO): Normal (95% CI) | 87.3%<br>(80.6 - 94.1) | 66.4%<br>(53.9 - 78.9) | 48.3%<br>(28.3 - 68.3) |
| ALT category (WHO): Elevated >ULN (95% CI) | 12.7%<br>(5.9 - 19.4) | 33.6%<br>(21.1 - 46.1) | 51.7%<br>(31.7 - 71.7) |

HIV: human immunodeficiency virus; HBV: hepatitis B virus; VL: viral load; SD: standard deviation; HBeAg: HBV e-antigen; ALT: alanine aminotransferase; CI - confidence intervals

For people receiving HIV ART, 4.9% of participants had HBV DNA VL > 2000 IU/mL with 1.3% (0.2% -8.7%) having VL 2001 - 20,000 IU/mL, while 3.6% (1.1% -11.7%) had VL > 20 000 IU/mL (**Table 3)**. For those not on ART, 18.2% had HBV DNA VL > 2000 IU/mL with 11.2% (6.5% -18.8%) being in the 2001 - 20,000 category, and 7.0% (3.8% -12.4%) were in the > 20 000 IU/mL category (**Table 3)**.

In spatial mapping, we observed heterogeneity in HBsAg prevalence, with the highest prevalence in the south and south-eastern region of the surveillance area, a peri-urban area, of which part borders a national highway. The lowest prevalences were concentrated in the more rural north-west and north (**Figure 4**).

**Figure 4.**
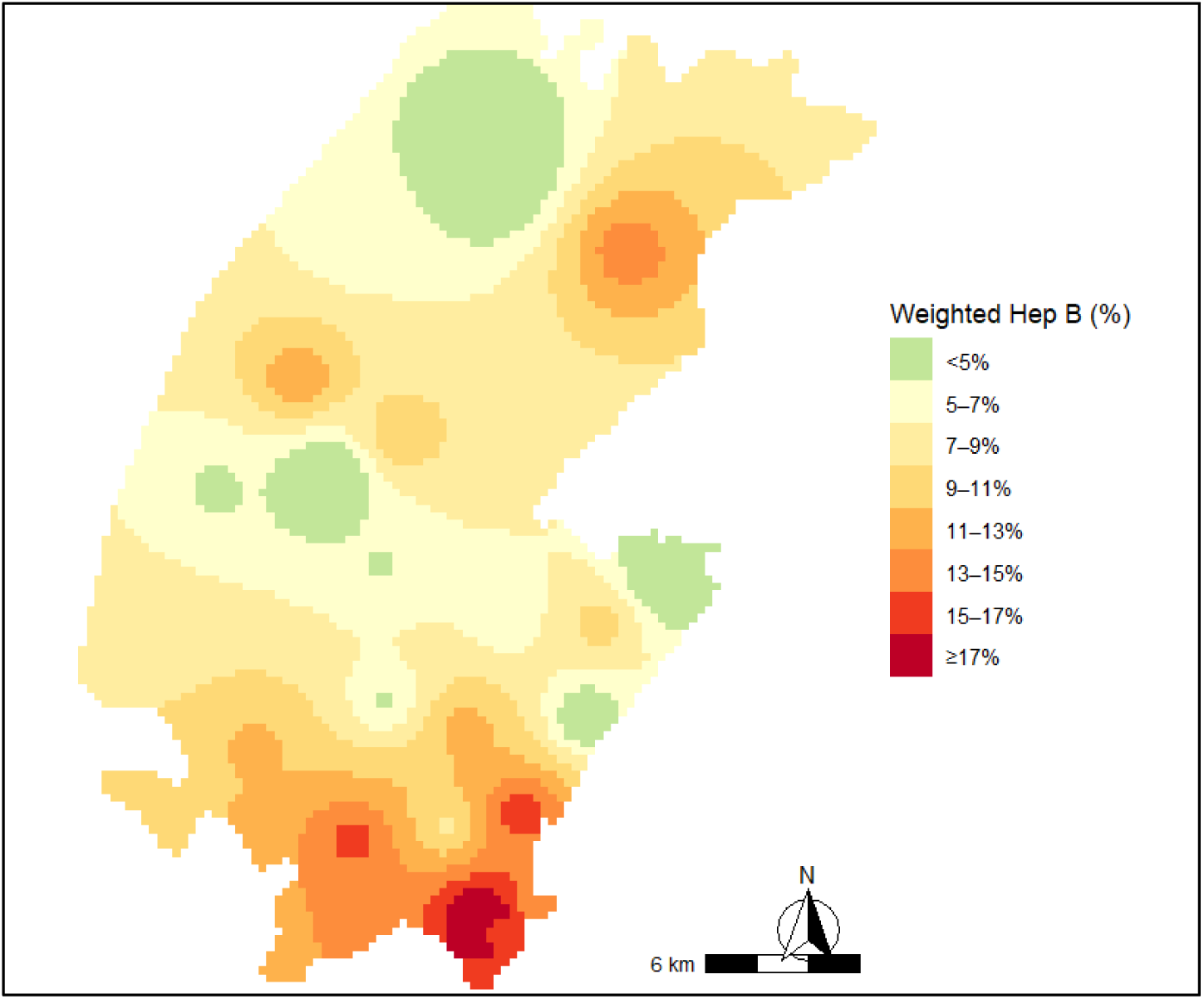
Spatial distribution of HBV prevalence in the uMkhanyakude district of rural Kwa-Zulu Natal, AHRI surveillance area. Continuous surface map of the southern part of the demographic surveillance area showing areas of lowest (green) and highest (red) prevalence for HBV based on HBsAg reported on Abbott assay (positive HBsAg S/CO >1). Prevalence is weighted to account for unequal probability of selection from the Vukazazi cohort into our study sample (by design), and for differential participation of the eligible population in the Vukuzazi study.

### 3.4 Factors associated with HBV immunity

#### (i) Vaccine mediated immunity

As expected, there were significant differences in the prevalence of vaccine-mediated HBV immunity between birth cohorts, reflecting immunisation of infants starting in the mid-1990’s. Among those born before 1995, vaccine-mediated immunity was only 5.5% (95% CI 4.1-7.2). Immunity related to vaccination was 13.0% (95% CI 9.8-17.0) for the 1995-1999 birth cohort, and 20.2% (95% CI 15.8-25.4) for those born between 2000-2005 (**Table 2**). This pattern remained after adjustment: compared with births before 1995, those born in 1995-1999 had 3.8 times higher odds of vaccine-mediated immunity (aOR 3.78, 95% CI 2.30-6.23) and those born in 2000-2005 had 6.1 times higher odds (aOR 6.12, 95% CI 3.52-10.64) (**Figure 3; Suppl Table 7).**

Vaccine-mediated immunity was lower in PLWH compared to HIV-negative individuals (6.7% (95% CI 5.0-8.9) vs 10.0% (95% CI 8.2–12.1); p=0.02) but this difference was not significant after adjustment (aOR 1.16, 95% CI 0.76-1.77; **Figure 3; Suppl Table 7**). Vaccine-mediated immunity was not statistically different between females vs males based on either crude estimates (9.1% (95% CI 7.3-11.3) vs 8.5% (95% CI 6.7-10.9) respectively) or after adjustment (aOR 1.07, 95% CI 0.72-1.58).

#### (ii) Clearance of natural infection

HBV exposure and clearance decreased sharply across birth cohorts (47.7%, 7.0%, 1.8%, for birth pre-1995, 1995-1999 and 2000 onwards, respectively; p<0.001; **Table 2**). This pattern remained significantly lower in younger cohorts in the adjusted model: compared to births before 1995, 1995-1999 had 91% lower odds (aOR 0.09, 95% CI 0.06-0.15) and 2000-2005 had 98% lower odds (aOR 0.02, 95% CI 0.01-0.05) (**Figure 3; Suppl Table 7**).

HBV exposure and clearance was higher in females on crude comparison (38.2% vs 30.1%; p=0.001), but this was not significant after adjustment (aOR 0.85, 95% CI 0.63-1.15; **Figure 3; Suppl Table 7**). Evidence of infection and clearance was more prevalent in PLWH (39.2%, 95% CI 35.1-43.3) than in HIV-negative individuals (27.3%, 95% CI 24.1-30.4) (**Table 4)** and this remained significant in the adjusted model (aOR 1.33, 95% CI 1.02-1.73; **Suppl Table 7**).

**Table 4:** Weighted prevalence of HBV vaccine mediated immunity and infection derived immunity in a rural population in Kwa-Zulu Natal, South Africa. Immunity is based on antiHBs titre >10 mIU/mL. Vaccine-derived immunity is defined as antiHBs in the absence of antiHBc. Infection-derived immunity is based on serological evidence of past infection (detectable antiHBc).

| Characteristic | Vaccine-mediated immunity<br>% (95% CI) | Infection-derived immunity<br>% (95% CI) |
| --- | --- | --- |
| Overall | 8.9 (7.5- 10.4) | 31.3 (28.8-33.8) |
| Sex |  |  |
| Female | 9.1 (7.3- 11.3) | 34.7 (31.1-38.3) |
| Male | 8.5 (6.7- 10.9) | 26.6 (23.5-29.7) |
| HBV vaccine age epoch |  |  |
| Born <1995 | 5.5 (4.1- 7.2) | 42.9 (39.6-46.1) |
| 1995-1999 | 13.0 (9.8- 17.0) | 6.1 (3.9-8.4) |
| 2000-2005 | 20.2 (15.8- 25.4) | 1.8 (0.2-3.4) |
| HIV status |  |  |
| Negative | 10.0 (8.2- 12.1) | 27.3 (24.1-30.4) |
| Positive | 6.7 (5.0- 8.9) | 39.2 (35.1-43.3) |

HIV: human immunodeficiency virus; HBV: hepatitis B virus;CI: confidence interval.

We also evaluated anti-HBs titres using different thresholds (**Suppl Figure 2 and Suppl Table 9)**. Based on a threshold of >100 mIU/mL, vaccine-mediated and infection-derived protection (anti-HBs positive) was present in 2.7% (95% CI 1.7-3.6) and 19.0% (95% CI 16.8-21.1), respectively. Overall, <0.1% of participants met the most stringent threshold for immunity of anti-HBs ≥1000 mIU/mL.

### 3.5 Susceptibility to HBV

Susceptibility was higher in younger, vaccine-era cohorts at 72.2% (born 1995-1999) and 71.4% (born 2000-2005) compared with 35.0% in those born pre-1995. Adjusted models confirmed this difference; compared to those born before 1995, participants born in 1995-1999 had 3.23 times the odds of being susceptible (aOR 3.23, 95% CI 2.38-4.37), and those born in 2000–2005 had 3.60 times the odds (aOR 3.60, 95% CI 2.51-5.15). Susceptibility was higher in males than females (50.1% vs 42.8% p=0.005, **Table 2**), although this crude sex difference attenuated after adjustment (**Figure 3**). Susceptibility was lower in PLWH than in those HIV-negative (35.4% vs 51.1%, respectively; p<0.001, **Table 2**), with a modelled estimate that PLWH had 36% lower odds of susceptibility than HIV-negative individuals (aOR 0.64, 95% CI 0.50-0.82). The reason for this difference in susceptibility is because there are higher rates of infection exposure in older adults, females and PLWH.

### 3.6 Relationship between HBV serostatus and alcohol, smoking, comorbidity, education and socioeconomic status

#### (i) Alcohol and smoking

Reporting drinking alcohol in the past 12 months was associated with lower odds of vaccine-mediated immunity compared with never-drinkers (aOR 0.49, 95% CI 0.25-0.95). Those reporting no alcohol in the last 12 months also had lower odds of HBV exposure and clearance (aOR 0.25, 95% CI 0.10-0.63), which is reflected in higher odds of being susceptible (aOR 2.92, 95% CI 1.48-5.77). There was no evidence of alcohol associations with HBV infection in adjusted models, and no evidence of association between smoking (current or previous) and any HBV seromarkers (**Suppl Table 7**).

#### (ii) Cardiovascular and metabolic risk factor

Diabetes and BMI were not associated with any serological pattern; **Suppl Table 7.** Hypertension was associated with lower odds of HBV susceptibility (aOR 0.70, 95% CI 0.50-0.98) but showed no association with infection, vaccine-mediated immunity, or exposure and clearance.

#### (iii) Education and socioeconomic status

Compared to those with no formal education, secondary and tertiary education were associated with lower odds of HBV exposure and clearance (secondary education, aOR 0.47, 95% CI 0.26-0.86; tertiary education aOR 0.47, 95% CI 0.25-0.89).

Mirroring this pattern, secondary education was associated with higher odds of susceptibility (aOR 1.98, 95% CI 1.17-3.35). There was no evidence of association between education and either infection or vaccine-mediated immunity; **Suppl Table 7.**

When comparing with the lowest SES, odds of exposure and clearance were increased at low, high and highest SES (low aOR 1.88, 95% CI 1.15-3.05; high aOR 2.75, 95% CI 1.64-4.63; highest aOR 2.27, 95% CI 1.35-3.81), while susceptibility was decreased at high and highest SES (high aOR 0.59, 95% CI 0.37-0.92; highest aOR 0.61, 95% CI 0.38-0.96). There was no association between SES and vaccine-mediated immunity. **Suppl Table 7.**

### 3.7 HBV treatment eligibility

Defining our population living with HBV infection using the primary Abbott assay threshold (HBsAg S/CO>1), 84/220 (38%) individuals were already receiving HBV-active antiviral treatment as a result of living with HIV coinfection (i.e ART). Among the remaining 156 individuals not on treatment, 14 met HBV treatment criteria based on HBV DNA >2000 and ALT >ULN. If also accounting for other comorbidity, a further 46 individuals could potentially be offered treatment (total population treated would thus be 60%; **Figure 5A**).

**Figure 5:**
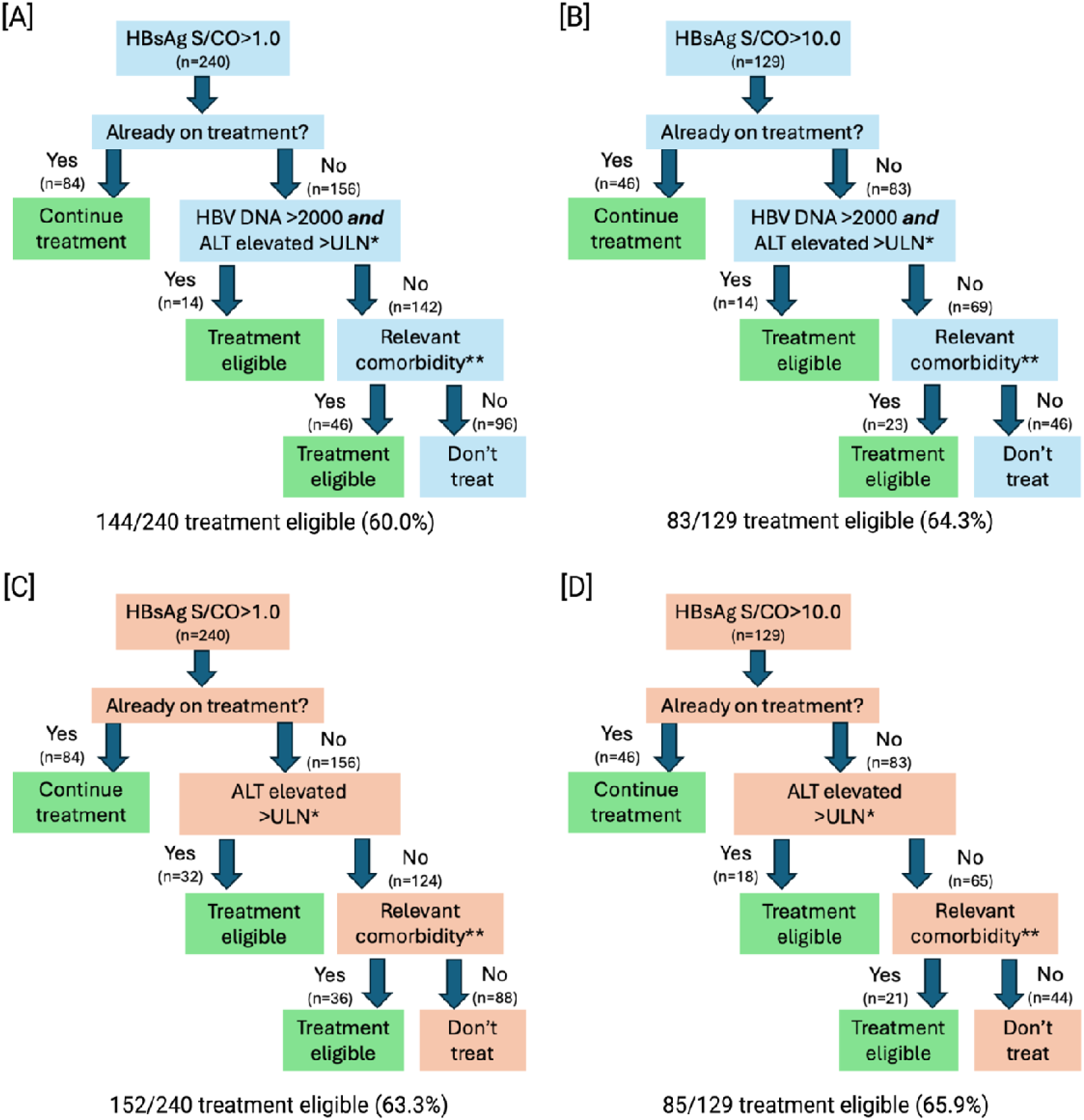
HBV treatment eligibility criteria for adults living with HBV infection in KwaZulu Natal, South Africa. [A] Assessment is based first on the relaxed HBV diagnostic criterion of HBsAg S/CO ratio >1.0 on Abbott assay with HBV DNA results available. [B] Assessment based on a more stringent HBsAg S/CO ratio of >10.0 with HBV DNA results available. [C] Assessment is based first on the relaxed HBV diagnostic criterion of HBsAg S/CO ratio >1.0 on Abbott assay but without accounting for HBV DNA quantification. [D] Assessment based on a more stringent HBsAg S/CO ratio of >10.0 but without accounting for HBV DNA quantification. ALT values categorised based on upper limit of normal (ULN) defined by WHO thresholds (30 U/L for males, 19 U/L for females). Relevant comorbidity is defined as a diagnosis of HIV or diabetes. * ALT above upper limit of normal (ULN) defined as 30 U/L for males and 19 U/L for females. ** HIV, diabetes.

Taking a more stringent approach to diagnosis, we reviewed 129 individuals with HBsAg S/CO >10, of whom 46 (26%) were on treatment as a result of HIV coinfection. Based on WHO laboratory criteria, an additional 14 became treatment eligible, and including those with comorbidity would suggest offering treatment to an additional 23 individuals (total population treated 64%; **Figure 5B**). As HBV DNA quantification is not universally available, and in line with 2024 WHO guidelines, we applied the same treatment algorithm using ALT as the only parameter. For HBsAg defined as S/CO >1.0, the total population treated was then 63%; (**Figure 5C**) and 66% for HBsAg >10.0 (**Figure 5D**).

## 4.0 DISCUSSION

### 4.1 Key findings

This study benefits from a unique and deeply characterized community-based, population-representative cohort with detailed metadata, and low missingness. Our data are important because there is no systematic approach to HBV screening in many African populations due to lack of funding, awareness, advocacy, and under-developed clinical and public health pathways. We highlight the significant ongoing public health threat associated with HBV in this region. Immunity is primarily driven by exposure, and while there is encouraging evidence of waning rates of exposure in younger populations, overall population immunity remains low (and therefore susceptibility is high), highlighting a pressing need for enhanced implementation of vaccination.

While HBV infection and exposure is commonly associated with an excess male risk^19^, we did not observe a sex-difference in this population, which may point to a relative vulnerability among women compared to other settings. The clustering and geographic variation highlight the non-random distribution of HBV infection across the study area, potentially pointing to local transmission dynamics or disparities in healthcare access. There is a high prevalence of NCDs, especially metabolic and cardiovascular disease, which can also contribute to chronic liver disease; this finding highlights that HBV should be tackled as part of syndemic population health needs.

### 4.2 Challenges in generating HBV prevalence estimates

We highlight challenges and inconsistencies that may arise in determination of HBsAg status, relevant both to diagnosis of individual infection and to generating accurate population-level estimates. Depending on the assay used, true positives may be missed (e.g. if testing relies on POCT), while more sensitive laboratory-based assays may be subject to false positive results, which can cause anxiety, impose costs on individuals and healthcare systems, and artificially inflate reported population prevalence estimates^16,17^. Typically, laboratory assays have been validated in Northern hemisphere populations, and there is limited understanding of performance metrics in African populations. Ideally, low-grade positive tests should be confirmed by an *in vitro* neutralisation assay^20^, with a previous study suggesting this should be applied at S/CO ratio between 1 and 25^21^; however, in practice this is not available in many settings and could not be undertaken within the scope of this study.

It is generally expected that the majority of HBsAg-positive samples would test anti-HBc positive, whereas in our study, only 53.8% of those with HBsAg S/CO >1.0 were reported anti-HBc positive. The same pattern has been reported in other studies, including from Zimbabwe, Botswana and Brazil^22^. This picture might be due to a false positive HBsAg, loss of anti-HBc over time, mutations which affect anti-HBc detection, an acute infection before anti-HBc is detectable, or immunosuppression including HIV infection^23^. In our study, we did not measure anti-HBc IgM which can be a marker of acute infection, but have assumed that the majority of those testing HBsAg-positive had chronic infection.

An alternative approach to ascertaining a true positive HBsAg test is to screen for HBV DNA. However, in this population, in which HIV and HBV are co-endemic, many individuals are receiving HBV-active antiviral drugs (either as treatment or prophylaxis for HIV); thus HBV DNA is not a reliable alternative screening tool. We conclude that, even with repeat testing, use of other serological markers, and/or testing HBV DNA, HBV status may remain equivocal.

Our approach to identification of HBV infection focused only on screening for HBsAg, and therefore did not identify cases of occult HBV infection (OBI), hence we might have underestimated HBV prevalence.

### 4.3 Groups at high risk

HIV and HBV share some epidemiological risk factors (e.g. injecting drug use or sexual exposure), which may explain the increased risk of HBV exposure we observed in PLWH. In this setting, alcohol was associated with lower vaccine-mediated immunity, although we did not find any relationship between this factor, or SES, with increased odds of HBV infection, in contrast to a prior study in a SA township^24^. However, as smoking and alcohol use were low in our population, we may have been underpowered to detect true associations. Differences may also be accounted for by varied HBV epidemiology between rural and urban settings^25^.

We explored the relationship between HBV infection and cardiometabolic risk factors given the high prevalence of these NCDs and an interest in the relationship between HBV and metabolic dysfunction associated steatotic liver disease (MASLD)^26^. There were no signals to suggest either risk or protection from HBV in association with hypertension, diabetes, overweight/obesity, but further work is needed to explore the risk-profiles and clinical features of those living with multiple risk factors for liver disease.

### 4.4 The future of HBV treatment

Based on WHO recommendations^3^, >60% of PLWHB in our study population met HBV treatment eligibility criteria. This reflects a high prevalence of HIV coinfection, but may underestimate the overall proportion with an indication for treatment, as we were not able to assess all eligibility criteria (e.g. hepatitis C status, steatosis, fibrosis and APRI scores^3^). These data highlight a high case of need for access to improved risk-assessment including diagnostic testing that is validated in different population settings, access to HBV DNA VL testing, and pathways for treatment assessment and provision in this setting and encourages quick adoption of the new recommended guidelines in order to avert disease progression.

Our previous research in this population has identified barriers to accessing healthcare interventions for HBV^27^. The impact of ART (including HBV-active agents) needs more investigation in this setting; widespread use of these agents may be contributing to the low HBV DNA VL we observed. At the time of this study, HBV treatment was only being accessed as a result of ART programmes for PLWH, with no access to therapy for those living with HBV monoinfection. Notably, only 72.5% of PLWH were on treatment, which may, in part, be due to some participants being newly diagnosed at the time of Vukuzazi enrolment and sample collection^15^. This coverage is lower than previously reported in Vukuzazi^18^, likely relating to our sample stratification (however, is higher than the 62.3% reported in a 2017 national population-based survey^28^). Thus, a reservoir of HBV and HIV persists in the population, which - combined with the low prevalence of vaccine-mediated immunity for HBV - is a risk factor for ongoing transmission.

Multipronged interventions are needed to tackle the ongoing public health challenge of co-endemic HBV and HIV, including education and advocacy, resources to support consistent implementation of the HBV-BD vaccine, access to vaccine programmes for infants (and older members of the population if they have risk factors for HBV infection), and provision of affordable, accessible diagnosis and antiviral treatment. Expanding access to HBV treatment will reduce the chances of individuals developing long-term liver disease, including HCC, while also providing population health benefits by reducing the risk of transmission. As highlighted by the WHO Global Hepatitis Report ‘Action for Access in Low- and Middle-Income Countries (LMICs)’, there is a need for decentralized services to reach those living in rural settings, and task-sharing to make delivery of services more efficient and accessible^29^.

New WHO guidelines^3^ also support use of dual therapy, which is more widely available as used as HIV ART, providing more accessible and cost-effective options in LMIC settings. The well-developed infrastructure for HIV service delivery in SA could be harnessed as an important platform for HBV education, testing, monitoring and treatment, but more implementation research is required to determine the optimum approaches to service delivery, especially as funding structures change.

### 4.5 HBV Immunity and informing vaccine programmes

In line with previous reports^6,30^, we observed a positive impact of the infant immunization programme in SA. However, while infant vaccine coverage for KZN has been reported as 75% in 2019^4^, our data suggest a lower rate of vaccine-mediated HBV immunity based on the prevalence of anti-HBs. In line with other studies in the KZN region^31^, we highlight that these populations are still far from the WHO targets for 90% vaccine coverage, and below the SA national estimates of 79%^32^. These data highlight inequity in vaccine coverage in SA (likely to be based on a combination of factors which include accessibility, education, acceptability, affordability and implementation). While renewed commitment by Gavi, the vaccine Alliance, promised resources to support HBV BD-vaccine programmes in some African settings, the changes in the funding landscape from early 2025 risk damaging already precarious vaccine delivery^33^.

Protective anti-HBs titres may not be observed, even after a full vaccine schedule. Waning immunity is recognized in PLWH^34^, and over time, which could also account for low prevalence of anti-HBs in this setting^30^. There are conflicting data regarding the relationship between chronic alcohol intake and HBV vaccine response, but as suggested in our study, alcohol may be associated with diminished immunity^35^, as well as being linked to other unmeasured SES factors which could affect outcomes.

### 4.6 Caveats and limitations

We adopted a cross-sectional approach to screening a limited number of stored samples. While we took a statistical approach to weighting to correct for the population sampled, this does not completely correct for bias in those who participated in the Vukuzazi programme, with a risk that the most vulnerable and marginalised individuals may be under-represented, and we did not include those age <15 years.

There were no HBV vaccination records available and our birth cohort definitions were based on the dates of introduction of infant vaccination in SA, but in real-world settings, access to vaccine is not consistent and vaccine programmes take time to embed; even in the vaccine epoch, not all those who are eligible receive a full course of HBV immunisation^6^. Self-reported variables, such as alcohol intake, can be influenced by how individuals interpret questions or what they feel comfortable disclosing. We might underestimate eligibility for therapy due to missingness of some eligibility markers however we also assumed HBV chronicity based on testing at one time point. Viral sequencing is proposed to better understand the molecular epidemiology of HBV in this setting, but in many samples is limited by low or undetectable HBV viraemia. It is proposed as a follow-up initiative, with the assessment of OBI which was also not included in this study. The generalisability of our results is uncertain, as the prevalence of co-endemic HIV, NCDs, access to laboratory testing and treatment pathways are highly setting-dependent.

### 4.7 Conclusions

Despite the roll-out of HBV vaccination since the mid-1990’s in SA, vaccine-mediated immunity is low and HBV infection and exposure in rural KZN remains endemic. Diagnostic and serological assays have different performance metrics and have not been robustly validated in African settings. Vulnerable groups may benefit from targeted interventions; for example, in KZN our data suggest a need to focus on women, those who drink alcohol, and people living in certain sub-regions where prevalence is high. Treatment-eligibility is often based on the presence of comorbidity, highlighting the need to break down disease-specific silos and focus on combined decentralised interventions, including enhanced education, access to diagnostic markers, and pathways to HBV treatment.

## Data Availability

All data produced in the present study are available upon reasonable request to the authors

## Acknowledgements

We are grateful to the participants of Vukuzazi, and the large AHRI clinical research team who supported this programme. We would like to acknowledge the Biostatistics Scientific Technology Platform (STP) at the Francis Crick Institute for contributions to statistical methods and discussion of analysis and presentation of data.

## Author contributions

Conceptualization: MA, TGM, PCM

Methodology: MA, LMa, TGM, KB, PCM Software: LMa, DG

Validation: NM, KB

Formal analysis: MA, LMa, SO, KB, PCM

Investigation: MA, GS, TGM, MD, EM, NM, LMt, MJS

Resources: GOJ, ME, RG, EBW, MJS, WAH, KH, TS

Data Curation: ME, GOJ, DG, KH

Management activities: TK, JU, TS, KH, WAH, PCM

Writing - Original Draft: MA, LMa, TGM, PCM

Writing - Review & Editing: all authors

Visualization: MA, LMa, KB, PCM

Supervision: VGN, TN, KB, CI, PCM

Project administration: EW, JU, TK

Funding acquisition: TGM, PCM

## Supplementary materials

File provided separately

