## supplemental material for "‘EVOLVE-HBV’: A retrospective cross-sectional study to quantify and characterise HBV infection, exposure, immunity and susceptibility in a rural population in KwaZulu-Natal, South Africa"

**CONTENTS OF SUPPLEMENT:**

**Supplementary methods	2**

Supplementary methods 1: Details of Vukuzazi programme	2

Supplementary methods 2: Stratified retesting of HBsAg	3

Supplementary methods 3: Statistical methods	4

Supplementary methods 4: Definitions and thresholds	6

Supplementary Results	7

Repeat HBsAg testing and sensitivity analysis for seroprevalence	7

Sensitivity analysis with changed diagnostic threshold for HBsAg	7

**Supplementary Tables	9**

Supplementary Table 1. Primary assays used to screen samples from individuals included in the EVOLVE HBV serosurvey from the Vukuzazi programme.	9

Supplementary Table 2. Additional diagnostic tests applied to a subset of samples testing HBsAg-positive on the primary assay platform.	10

Supplementary Table 3: Summary of 2024 WHO HBV Treatment Criteria and application to EVOLVE-HBV study cohort	11

Supplementary Table 4: Attributes of assay performance for HBsAg testing (based on outcomes in KZN population)	12

Supplementary Table 5: Weighted HBsAg seroprevalence calculated based on altered HBsAg S/CO threshold applied to the Abbott (Architect) diagnostic assay.	13

Supplementary Table 6. Survey-weighted logistic regression for HBsAg positive status based on S/CO >1 and S/CO >10, adjusted for sex, birth epoch and HIV status.	15

Supplementary Table 7. Predictors of HBV exposure and clearance, vaccine mediated immunity and susceptibility	16

Supplementary Table 8. Predictors of HBV exposure and clearance, vaccine mediated immunity and susceptibility at HBsAg S/CO<10	18

**Supplementary Figures	21**

Supplementary Figure 1: Distribution of results of HBsAg S/CO >1.0 on Abbott assay.	21

Supplementary Figure 2. Distribution of anti-HBs titres in 2200 adults sampled from the Vukuzazi cohort in KwaZulu Natal, South Africa.	23

Supplementary Figure 3. Forest plot of multivariable analysis of factors associated with HBV serostatus.	24

Supplementary References	25

### **Supplementary methods**

#### Supplementary methods 1: Details of Vukuzazi programme

Starting in 2018, Vukuzazi built on a local public health system, gathering demographic and health information to survey the community's health using mobile clinics, collect data, screen and refer for treatment, and obtain a broad range of population-based data and biological samples. Vukuzazi participation was offered to resident members of households in the AHRI Health and Demographic Surveillance Systems (HDSS) area, who were aged ≥15 years between May 2018 and March 2020. As a result of a highly structured approach to prospective data collection within the Vukuzazi programme, there was hardly any missing data in our study sample (the highest missingness was 3% on the SES variable). Plasma samples collected by the Vukuzazi programme were stored in aliquots of 500 ul at -80°C in a biorepository at AHRI, Durban.

We collated information from the Vukuzazi dataset including alcohol and tobacco use, socio economic status (SES) quintiles , ART, HIV testing and HIV VL, BMI, and blood pressure.

In this study, our pre-vaccine birth cohort comprises individuals born 1985-1994 (n=303), born 1975-1984 (n=300), born 1965-1974 (n=300), and born before 1965 (n=300).

The Vukuzazi programme was approved by the UKZN Biomedical Research Ethics Committee (BREC) (ref. BE560/17), the London School of Hygiene & Tropical Medicine (LSHTM) (ref. 14722), the Partners Institutional Review Board (ref. 2018P001802), and the University of Alabama at Birmingham (ref. 300007237).

#### Supplementary methods 2: Stratified retesting of HBsAg

In clinical practice, equivocal HBsAg test results are recognised, and different assays have varying performance metrics. Repeat testing may be recommended for samples at the lower end of the HBsAg sample/cut-off (S/CO) range. Our study protocol did not include formal assessment of assay metrics or validation of diagnostic testing. However, to better understand assay performance, we undertook repeat testing of 20 samples reported HBsAg-positive by our primary laboratory assay (Abbott Architect), using both a point-of-care test and an alternative validated laboratory platform to investigate concordance between different diagnostic methods. The 20 samples were chosen to reflect a range of HBsAg S/CO ratios determined by the initial assay (range 1.07-5101.18).

#### Supplementary methods 3: Statistical methods

This was an exploratory study with the primary aim of estimating the prevalence of HBV biomarkers. As such, the sample size was based on feasibility and budget considerations, rather than formal power calculations. Based on existing estimates of 4-8% prevalence of HBV infection in the SA population^1–8^, with a sample size of 2200, we would be able to estimate this prevalence with a precision of 0.9-1.2% with 95% confidence (e.g. if point estimate was 8%, the 95% confidence interval would be 6.8-9.2%). With this sample size, we would expect to find between 88-176 cases of HBV infection.

Our study sample of 2200 Vukuzazi participants was stratified by birth cohort, sex and HIV status. We applied weights to account for the unequal probability of selection from the Vukazazi cohort into our study sample (by design), and for differential participation (non-response) of the eligible HDSS population in the Vukuzazi cohort. The weighting was used to account for under- and over-representation of sex, HIV status and age groups, so that our sample more closely represented the population from which it was drawn. Sampling weights were calculated as the inverse probability of selection into our study in strata defined by birth cohort, sex, and HIV status. Non-response weights were calculated as the inverse probability of participation in Vukuzazi, using a logistic regression model including all eligible individuals in the HDSS area (captured through household surveys 3 times a year), with covariates for age group (<25, 25-44, 45-64 and ≥65 years) and sex; the inverse of the predicted probability from the regression model was used as the weight. Final weights ranged from 1.85 to 35.4, with 10% of observations having weights <2.3 and 10% with weights >28.4; no trimming was applied.

Confidence intervals for weighted prevalences were calculated using a logit transformation, with Taylor series linearisation variance estimation. Although sampling was random within age-sex-HIV strata, 12.6% of samples were from participants who lived in the same household. Clustering within household was not taken into account in the analyses; however, the estimated design effect ranged from 1.44 (vaccine-mediated immunity) to 1.60 (HepB antibody).

In the adjusted analyses of factors associated with the 3 HBV outcomes (HBV infection, HBV exposure and clearance, and vaccine mediated immunity), we included the stratification variables (birth cohort, sex and HIV status) and education, SES, alcohol intake, smoking status, BMI category, HTN and diabetes. All variables were retained in the adjusted model, irrespective of whether they remained significantly associated with the outcome at p<0.05 after adjustment.

We did not do multiple imputation for missing data as the outcomes and main exposures of interest were complete; the covariate with the highest proportion of missing data (SES, 3%) was not of primary interest for this study. This was an exploratory study and no adjustments were made for multiple testing; therefore, because of the potential for type I error, findings should be interpreted with caution.

#### Supplementary methods 4: Definitions and thresholds

Socioeconomic status (SES) was measured using a household asset index derived from ownership of durable goods, housing materials, and utilities. Principal components analysis (PCA) was used to generate a composite SES score for all households in the HDSS, which was then divided into quintiles, with the first quintile representing the most socioeconomically deprived households and the fifth quintile representing the least deprived.

HBV viral suppression was defined as VL <10 IU/mL. HIV VL suppression was defined as <50 copies/mL. Anti-HBs levels ≥10 mIU/mL were considered positive/protective.

Body Mass Index (BMI) categories were defined as underweight (<18.5 kg/m²); normal (18.5 - 24 kg/m²), overweight (25 - 30 kg/m²) and obese (>30 kg/m²).

Hypertension (HTN) was defined as systolic blood pressure ≥140mmHg or diastolic blood pressure ≥90 mmHg.

ALT values were categorised using the upper limit of normal (ULN) as defined by the World Health Organization (30 U/L for males, 19 U/L for females).

### **Supplementary Results**

#### Repeat HBsAg testing and sensitivity analysis for seroprevalence

We retested 20 samples reported as HBsAg on the index assay, among which only five were reported as positive by POCT, all of which had high HBsAg S/CO ratios reported by Abbott (**Suppl Figure 1A,B,C**). A second diagnostic assay, Cobas, was applied to 19/20 samples (one had insufficient volume); results were concordant with the POCT. These findings reflect a lower sensitivity of these alternative approaches for detecting HBsAg, but also identify potentially low specificity of our primary screening method (**Suppl Table 4**).

Among 15 samples testing positive on the Abbott assay, but negative on POCT (±Cobas assay), 7/15 had either HBV DNA+ or other serological evidence of HBV infection, while the other 8 remain equivocal (**Suppl Figure 1B**).

We explored the impact of applying more stringent HBsAg S/CO thresholds among 240 samples that were originally reported positive. Using stricter positivity definitions (higher S/CO threshold), weighted HBV seroprevalence declined (**Suppl** **Table 5**); at S/CO >10, HBV prevalence was 5.9% (95% CI 4.9-6.9%) (**Suppl** **Table 5**).

#### Sensitivity analysis with changed diagnostic threshold for HBsAg

Using the more stringent HBsAg positivity threshold of S/CO>10, HBV prevalence was 8.4% (6.8-10.0) in those born before 1995, 3.7% (2.2-5.3) in 1995-1999, and 1.6% (0.4-2.8) in 2000-2005 (p<0.001). Even in the adjusted model, HBV infection odds were lower in younger cohorts and statistically significant. Compared to those born before 1995, the 1995-1999 had 52% lower odds of infection (aOR 0.48, 95% CI 0.24-0.96; and among 2000-2005, the odds of HBV infection were 87% lower (aOR 0.13, 95% CI 0.03-0.46). By sex, prevalence was 4.4% (95% CI 3.3-5.6) in females and 7.6% ( 95% CI 6.0-9.3) in males; the adjusted estimate was consistent with lower odds in females, though not statistically significant (aOR 0.64, 95% CI 0.37-1.09). Likewise HIV status showed no evidence of association at this threshold: prevalence of HBV infection was 6.6% (95% CI 5.1-8.0) in PLWH vs 5.2% (95% CU 3.9-6.5) in HIV-negative, with aOR 1.33 (95% CI 0.82-2.18). **Suppl** **Tables 6-8; supplementary Figure 3 .**

Again based on HBV infection defined as HBsAg S/CO>10, compared to those born before 1995, the 1995-1999 had 52% lower odds of infection (aOR 0.48, 95% CI 0.24-0.96; and among 2000-2005, the odds of HBV infection were 87% lower (aOR 0.13, 95% CI 0.03-0.46). **Suppl** **Tables 6-8; supplementary Figure 3 .**

### **Supplementary Tables**

#### Supplementary Table 1. Primary assays used to screen samples from individuals included in the EVOLVE HBV serosurvey from the Vukuzazi programme.

| **Laboratory**  **test** | **Type of output** | **Platform** | **Limits of detection or cut-off threshold** | **Limit of quantification** |
| --- | --- | --- | --- | --- |
| HBsAg | Categorical | Abbott Architect System (Abbott Park, Illinois, USA) | S/CO ratio 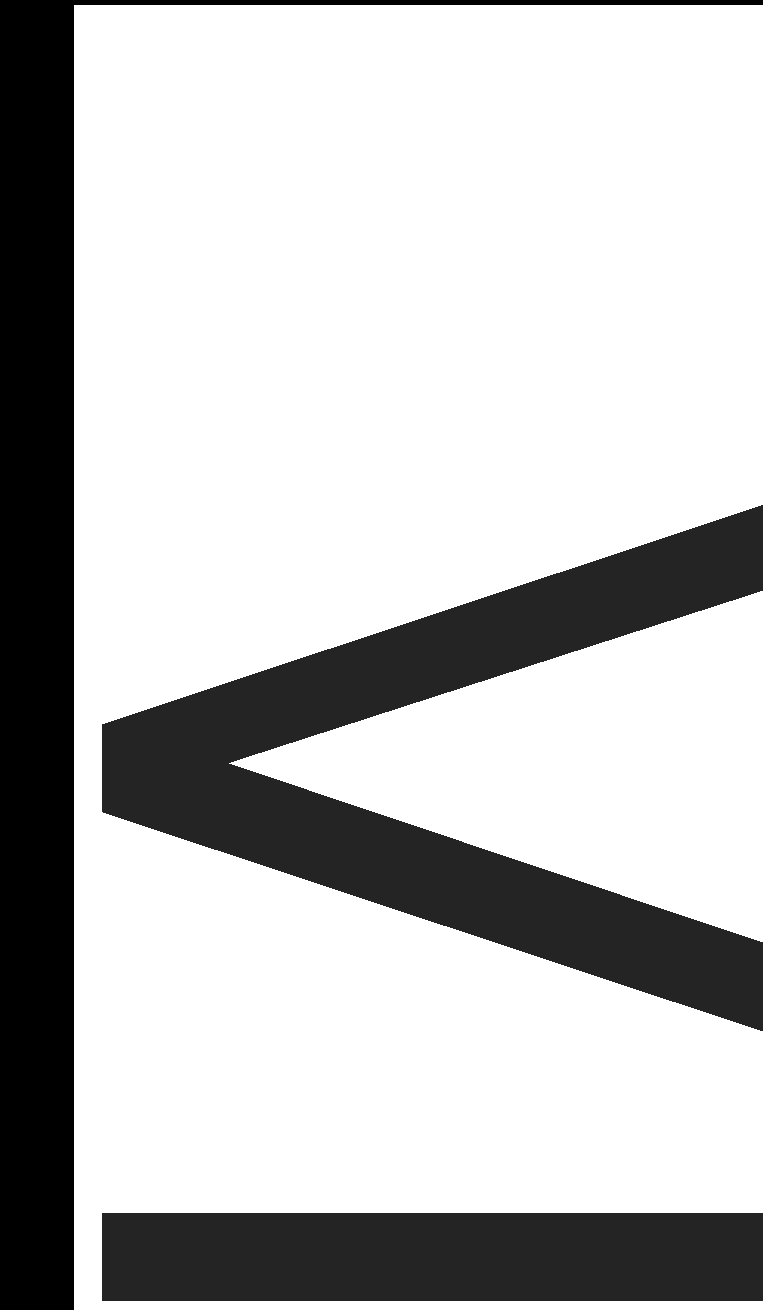1 negative  S/CO ratio >1 positive | 0.05 IU/mL |
| Anti-HBc | Qualitative | Abbott Architect System  Abbott Park, Illinois, USA) | S/CO ratio 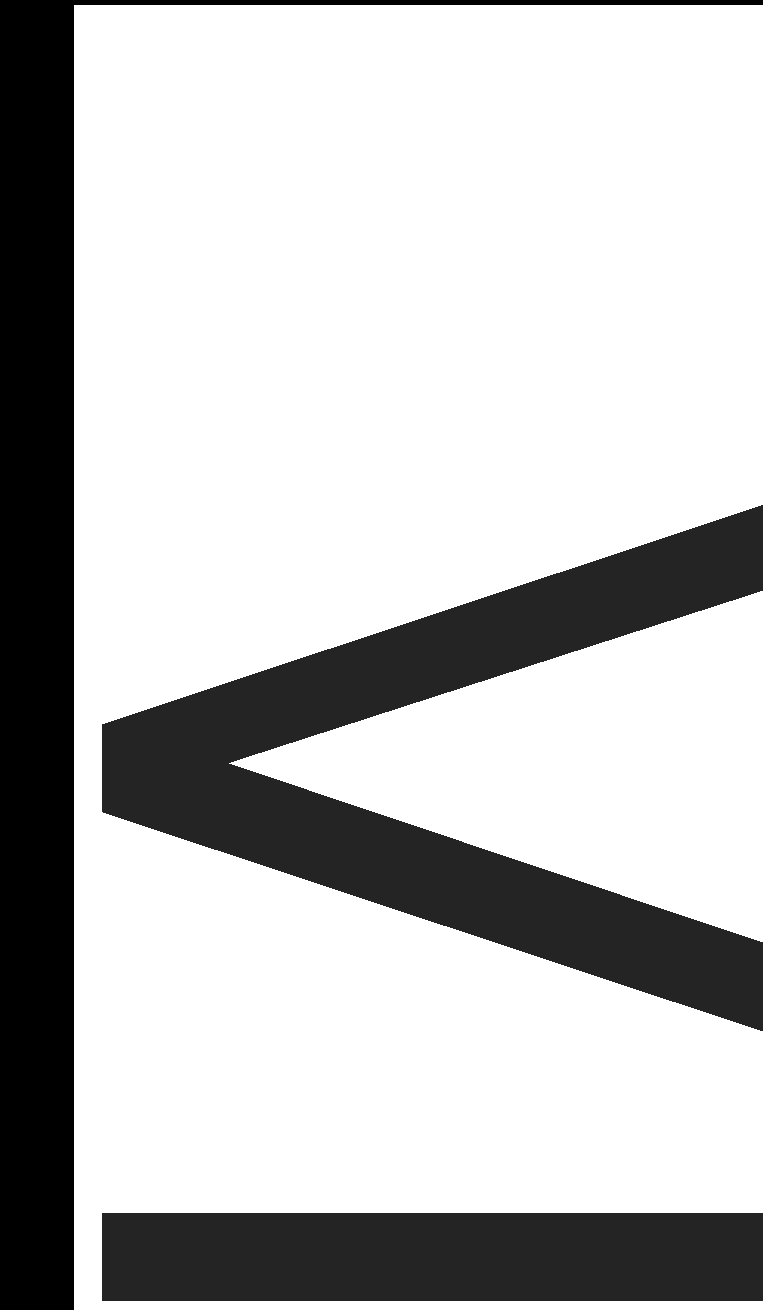1 negative  S/CO ratio >1 positive | NA |
| HBeAg* | Qualitative | Abbott Architect System (Abbott Park, Illinois, USA) | S/CO ratio 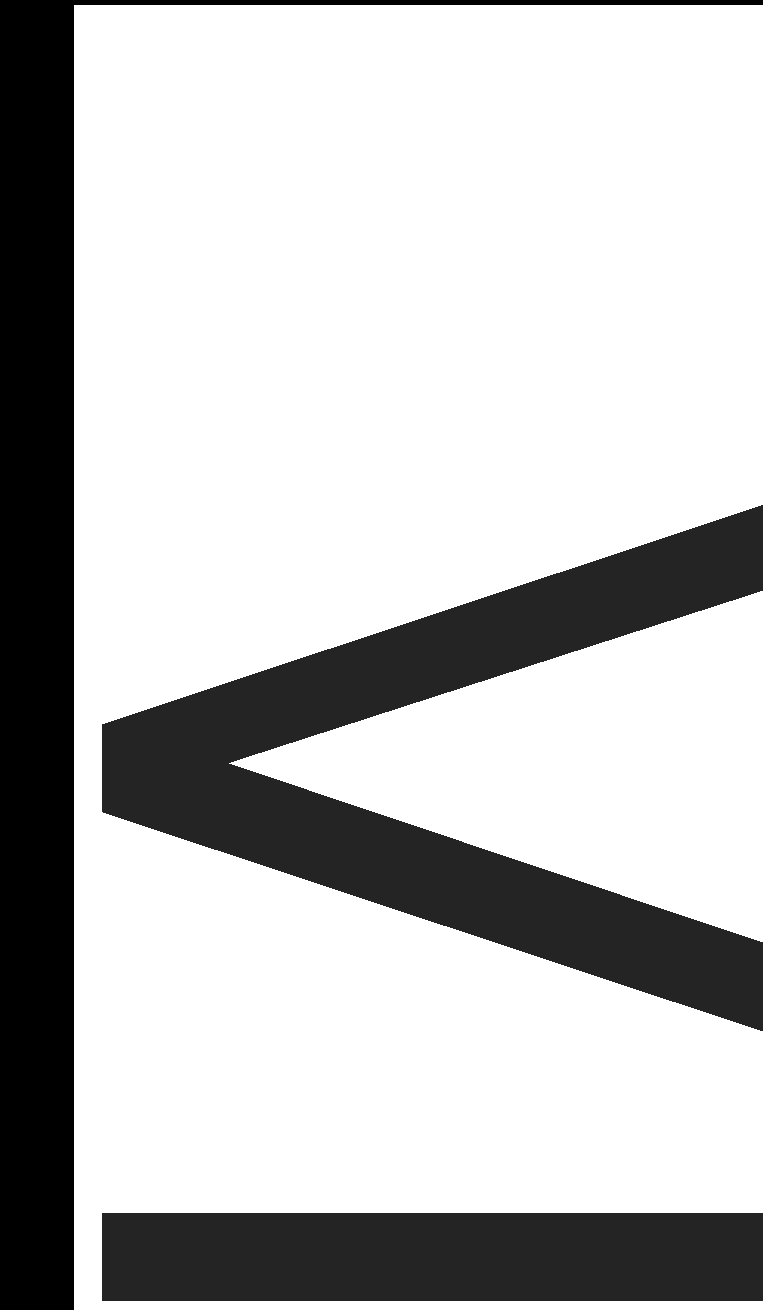1 negative  S/CO ratio >1 positive | NA |
| Anti-HBs | Quantitative | Abbott Architect System  Abbott Park, Illinois, USA) | LLoD 2.5 mIU/mL | ULoQ 1000 mIU/mL |
| HBV viral load* | Quantitative | Abbott Alinity m (Abbott Park, Illinois, USA) | LLoD 4.29 IU/mL | 10 IU/mL-1x10^9^ IU/mL |
| ALT* | Quantitative | Siemens Atellica (Siemens Healthineers, USA) | NA | NA |

* Test conducted only in 240 samples which tested HBsAg-positive on Abbott assay. HBV: Hepatitis B Virus; anti-HBc: HBV core antibodies; HBsAg: HBV surface antigen; anti-HBs: HBV surface antibodies; ALT: Alanine Aminotransferase; NA: Not applicable; LLoD: Lower limit of detection; ULoQ: Upper limit of quantification; S/CO: sample/cut-off. Chemiluminescence assays were performed at the National Health Laboratory Service (NHLS) in Durban. Positive and negative controls were included in each run to detect contamination and identify potential false positive / negative results.

#### Supplementary Table 2. Additional diagnostic tests applied to a subset of samples testing HBsAg-positive on the primary assay platform.

| **Laboratory**  **test** | **Type of output** | **Platform** | **Limits of detection or cut-off threshold** | **Limit of quantification** |
| --- | --- | --- | --- | --- |
| HBsAg | Categorical | CobasPro e801 * (Roche Diagnostics, Mannheim, Germany) | S/CO ratio ≤1 negative  S/CO ratio >1 positive | NA |
| HBsAg | Categorical | Determine™ HBsAg 2 (Abbott Park, Illinois, USA) POCT** | Cutoff index (signal sample/cutoff)  0.9 (0.020 IU/mL) | NA |

* Cobas assay performed at Lancet laboratories, Durban, South Africa

** POCT - point of care test - performed at Africa Health Research Institute

HBsAg: hepatitis B surface antigen; S/CO: sample/cut-off.

NA - not applicable

#### Supplementary Table 3: Summary of 2024 WHO HBV Treatment Criteria and application to EVOLVE-HBV study cohort

| **WHO Treatment Criteria (2024 Guidelines)** | **Description** | **Application in Current Study** |
| --- | --- | --- |
| 1. HBV DNA >2000 IU/mL and ALT > ULN | Antiviral therapy recommended if both HBV DNA exceeds 2000 IU/mL and ALT is above the upper limit of normal (ULN). | Applied as per guideline when both values were available. |
| 2. Co-infections or Comorbidities | Treatment recommended for individuals with co-infections (e.g. HIV) or comorbidities (e.g. diabetes), regardless of HBV DNA or ALT levels. | Applied as per guideline. |
| 3. ALT Persistently > ULN (when HBV DNA unavailable) | Treatment recommended if ALT is persistently >ULN (two measurements over 6-12 months) in settings without HBV DNA testing. | Modified: single ALT >ULN result considered due to the cross-sectional study design. |
| 4. Advanced Liver Disease (Fibrosis/Cirrhosis) | Treatment recommended based on imaging (e.g. transient elastography) or laboratory markers (e.g. APRI score). | Not applied — transient elastography and APRI data were not available in this study. |

HBV: hepatitis B virus; ALT: Alanine Aminotransferase; APRI: Aspartate transaminase to Platelet Ratio Index

#### Supplementary Table 4: Attributes of assay performance for HBsAg testing (based on outcomes in KZN population)

| **Abbott** | **Cobas*** | **POCT*** |
| --- | --- | --- |
| - Wide range of attributes of samples testing HBsAg-positive, including antiHBs and antiHBc status, HBV viraemia - Highly sensitive down to very low S/CO ratio but therefore at risk of calling false positives (less specific than other assays) | - Detects only the samples with high HBsAg S/CO (lower limit based on this sample set ~4500), ie limited sensitivity - Among Cobas+ samples, majority are viraemic (4/5), and all are anti-HBs negative and anti-HBc positive - All that are Cobas+ are also POCT+ | - Detects only the samples with high HBsAg S/CO (lower limit based on this sample set ~4300), ie limited sensitivity - Among POCT+ samples, majority are viraemic (4/5), and all are anti-HBs negative and anti-HBc positive - All that are POCT+ are also Cobas+ (though one not retested on Cobas) |

* Assessment limited by small numbers tested in this study, and all samples tested by Cobas / POCT were reported as positive by Abbott at baseline.

HBsAg: hepatitis B surface antigen; S/CO: sample/cut-off; HBV: hepatitis B virus; POCT: point of care test

#### Supplementary Table 5: Weighted HBsAg seroprevalence calculated based on altered HBsAg S/CO threshold applied to the Abbott (Architect) diagnostic assay.

| **Threshold (S/CO > k)** | **Characteristic** | **Sub-group** | **Prevalence % (95% CI)** | **p-value** |
| --- | --- | --- | --- | --- |
| >2 | Overall | All | 8.8 (7.6-10.0) | - |
|  | Birth Epoch | Born <1995 | 11.6 (9.8-13.5) | <0.001 |
|  |  | 1995-1999 | 5.2 (3.3-7.0) |  |
|  |  | 2000-2005 | 5.5 (3.4-7.7) |  |
|  | Sex | Female | 7.1 (5.7-8.6) | 0.003 |
|  |  | Male | 10.8 (8.8-12.7) |  |
|  | HIV status | HIV-negative | 7.7 (6.2-9.3) | 0.083 |
|  |  | Living with HIV | 9.8 (8.1-11.6) |  |
| >5 | Overall | All | 6.6 (5.5-7.6) | - |
|  | Birth Epoch | Born <1995 | 9.4 (7.7-11.0) | <0.001 |
|  |  | 1995-1999 | 3.7 (2.2-5.3) |  |
|  |  | 2000-2005 | 2.3 (0.9-3.7) |  |
|  | Sex | Female | 5.4 (4.2-6.7) | 0.021 |
|  |  | Male | 7.9 (6.2-9.6) |  |
|  | HIV status | HIV-negative | 5.7 (4.4-7.1) | 0.121 |
|  |  | Living with HIV | 7.4 (5.8-8.9) |  |
| >10 | Overall | All | 5.9 (4.9-6.9) | - |
|  | Birth Epoch | Born <1995 | 8.4 (6.8-10.0) | <0.001 |
|  |  | 1995-1999 | 3.7 (2.2-5.3) |  |
|  |  | 2000-2005 | 1.6 (0.4-2.8) |  |
|  | Sex | Female | 4.4 (3.3-5.6) | 0.293 |
|  |  | Male | 7.6 (6.0-9.3) |  |
|  | HIV status | HIV-negative | 5.2 (3.9-6.5) | 0.038 |
|  |  | Living with HIV | 6.6 (5.1-8.0) |  |
| >100 | Overall | All | 4.6 (3.7-5.5) | - |
|  | Birth Epoch | Born <1995 | 7.0 (5.5-8.4) |  |
|  |  | 1995-1999 | 2.5 (1.2-3.8) | <0.001 |
|  |  | 2000-2005 | 0.7 (−0.1-1.5) | <0.001 |
|  | Sex | Female | 3.0 (2.1-4.0) | <0.001 |
|  |  | Male | 6.5 (5.0-8.1) |  |
|  | HIV status | HIV-negative | 4.1 (2.9-5.3) | 0.262 |
|  |  | Living with HIV | 5.1 (3.8-6.4) |  |

*Notes:* Survey-weighted proportions with 95% CIs; strict thresholds (S/CO > k ⇒ S/CO ≤ k is negative). P-values are design-based tests for differences within each stratum vs. the reference category (Born <1995; HIV-negative; female). HIV: human immunodeficiency virus.

#### Supplementary Table 6. Survey-weighted logistic regression for HBsAg positive status based on S/CO >1 and S/CO >10, adjusted for sex, birth epoch and HIV status.

| **Predictor** | **HBsAg+ (S/CO>1)** | **HBsAg+ (S/CO>10)** |
| --- | --- | --- |
| **HBV vaccine age epoch** | | |
| Born <1995 | 1 (Reference) | 1 (Reference) |
| 1995-1999 | 0.65 (0.42-1.02) | 0.40 (0.21-0.75) |
| 2000-2005 | 0.54 (0.32-0.91) | 0.14 (0.05-0.41) |
| **Sex** | | |
| Male | 1 (Reference) | 1 (Reference) |
| Female | 0.76 (0.55-1.06) | 0.65 (0.41-1.02) |
| **HIV status** | | |
| HIV-negative | 1 (Reference) | 1 (Reference) |
| PLWH | 1.30 (0.92-1.85) | 1.24 (0.78-1.97) |

HBsAg: hepatitis B surface antigen; S/CO: sample/cut-off; HBV: hepatitis B virus; HIV: human immunodeficiency virus; PLWH – person living with HIV.

#### Supplementary Table 7. Predictors of HBV exposure and clearance, vaccine mediated immunity and susceptibility

Odds ratio with 95% CI are presented. These data are also plotted in Figure 3.

| Predictor | HBV infection (HBsAg S/CO>1) | HBV infection (HBsAg S/CO≥10) | Vaccine-mediated immunity  (S/CO>1) | HBV exposure & clearance  (S/CO>1) | Susceptible  (S/CO>1) |
| --- | --- | --- | --- | --- | --- |
| HBV vaccine age epoch | | | | | |
| Born <1995 | 1 (Reference) | 1 (Reference) | 1 (Reference) | 1 (Reference) | 1 (Reference) |
| 1995-1999 | 0.79 (0.48-1.29) | 0.48 (0.24-0.96) | 3.78 (2.30-6.23) | 0.09 (0.06-0.15) | 3.23 (2.38-4.37) |
| 2000-2005 | 0.69 (0.37-1.28) | 0.13 (0.03-0.46) | 6.12 (3.52-10.64) | 0.02 (0.01-0.05) | 3.60 (2.51-5.15) |
| Sex | | | | | |
| Male | 1 (Reference) | 1 (Reference) | 1 (Reference) | 1 (Reference) | 1 (Reference) |
| Female | 0.78 (0.52-1.19) | 0.64 (0.37-1.09) | 1.07 (0.72-1.58) | 0.85 (0.63-1.15) | 1.21 (0.93-1.58) |
| Education | | | | | |
| No formal | 1 (Reference) | 1 (Reference) | 1 (Reference) | 1 (Reference) | 1 (Reference) |
| Primary | 1.05 (0.47-2.38) | 0.45 (0.18-1.17) | 1.06 (0.37-3.07) | 0.98 (0.50-1.91) | 0.96 (0.53-1.73) |
| Secondary | 1.12 (0.53-2.41) | 0.77 (0.30-1.98) | 0.76 (0.29-1.95) | 0.47 (0.26-0.86) | 1.98 (1.17-3.35) |
| Tertiary | 1.52 (0.67-3.46) | 1.20 (0.44-3.33) | 0.93 (0.33-2.68) | 0.47 (0.25-0.89) | 1.63 (0.91-2.91) |
| SES | | | | | |
| Lowest | 1 (Reference) | 1 (Reference) | 1 (Reference) | 1 (Reference) | 1 (Reference) |
| Low | 1.07 (0.59-1.94) | 0.84 (0.38-1.83) | 0.61 (0.30-1.21) | 1.88 (1.15-3.05) | 0.70 (0.46-1.07) |
| Middle | 0.94 (0.51-1.75) | 0.74 (0.33-1.68) | 0.86 (0.45-1.65) | 1.47 (0.89-2.41) | 0.82 (0.53-1.26) |
| High | 0.82 (0.43-1.57) | 0.73 (0.31-1.70) | 0.56 (0.27-1.16) | 2.75 (1.64-4.63) | 0.59 (0.37-0.92) |
| Highest | 0.88 (0.46-1.70) | 0.65 (0.27-1.57) | 0.80 (0.41-1.59) | 2.27 (1.35-3.81) | 0.61 (0.38-0.96) |
| Alcohol intake | | | | | |
| Never drinker | 1 (Reference) | 1 (Reference) | 1 (Reference) | 1 (Reference) | 1 (Reference) |
| No drinking in last 12 months | 0.63 (0.21-1.88) | 1.12 (0.33-3.81) | 1.14 (0.32-4.03) | 0.25 (0.10-0.63) | 2.92 (1.48-5.77) |
| Drinking in past 12 months | 1.09 (0.62-1.89) | 1.01 (0.49-2.04) | 0.49 (0.25-0.95) | 0.91 (0.61-1.36) | 1.20 (0.83-1.72) |
| Smoking | | | | | |
| Never | 1 (Reference) | 1 (Reference) | 1 (Reference) | 1 (Reference) | 1 (Reference) |
| Ever smoked (current/former) | 1.30 (0.74-2.28) | 0.99 (0.51-1.94) | 0.79 (0.37-1.68) | 0.95 (0.61-1.48) | 0.95 (0.62-1.45) |
| HIV status | | | | | |
| Negative | 1 (Reference) | 1 (Reference) | 1 (Reference) | 1 (Reference) | 1 (Reference) |
| Positive | 1.36 (0.95-1.96) | 1.33 (0.82-2.18) | 1.16 (0.76-1.77) | 1.33 (1.02-1.73) | 0.64 (0.50-0.82) |
| BMI category | | | | | |
| Normal | 1 (Reference) | 1 (Reference) | 1 (Reference) | 1 (Reference) | 1 (Reference) |
| Underweight | 0.88 (0.40-1.95) | 1.58 (0.59-4.22) | 0.42 (0.18-0.97) | 1.61 (0.85-3.04) | 1.07 (0.68-1.69) |
| Overweight | 1.14 (0.73-1.76) | 0.91 (0.51-1.62) | 1.07 (0.66-1.74) | 1.21 (0.86-1.71) | 0.78 (0.57-1.07) |
| Obese | 1.19 (0.72-1.98) | 1.02 (0.55-1.89) | 1.23 (0.73-2.08) | 1.06 (0.73-1.53) | 0.84 (0.59-1.19) |
| Hypertension | | | | | |
| No | 1 (Reference) | 1 (Reference) | 1 (Reference) | 1 (Reference) | 1 (Reference) |
| Yes | 1.08 (0.68-1.72) | 1.44 (0.81-2.58) | 1.09 (0.55-2.14) | 1.29 (0.92-1.79) | 0.70 (0.50-0.98) |
| Diabetes | | | | | |
| No | 1 (Reference) | 1 (Reference) | 1 (Reference) | 1 (Reference) | 1 (Reference) |
| Yes | 1.17 (0.62-2.18) | 1.32 (0.61-2.89) | 1.68 (0.75-3.74) | 1.00 (0.61-1.64) | 0.79 (0.47-1.33) |

HBV: hepatitis B virus; HIV: human immunodeficiency virus; Body Mass Index: BMI; SES - socioeconomic status

#### Supplementary Table 8. Predictors of HBV exposure and clearance, vaccine mediated immunity and susceptibility at HBsAg S/CO<10

Odds ratio with 95% CI are presented. These data are also plotted in supplementary Figure 3.

| Predictor | Vaccine-mediated immunity  (S/CO<10; anti-HBs+; anti-HBc−) | HBV exposure & clearance  (S/CO<10; anti-HBc+) | Susceptible  (S/CO<10; anti-HBc−; anti-HBs−) |
| --- | --- | --- | --- |
| Age at enrolment (years) | 1.02 (1.00–1.05) | 1.04 (1.03–1.06) | 0.95 (0.94–0.96) |
| HBV vaccine age epoch |  |  |  |
| Born <1995 | 1 (Reference) | 1 (Reference) | 1 (Reference) |
| 1995–1999 | 6.46 (3.42–12.18) | 0.19 (0.12–0.31) | 1.50 (1.03–2.18) |
| 2000–2005 | 12.03 (5.62–25.78) | 0.07 (0.03–0.17) | 1.18 (0.73–1.91) |
| Sex |  |  |  |
| Male | 1 (Reference) | 1 (Reference) | 1 (Reference) |
| Female | 1.06 (0.72–1.56) | 0.88 (0.64–1.20) | 1.21 (0.91–1.59) |
| Education level |  |  |  |
| None | 1 (Reference) | 1 (Reference) | 1 (Reference) |
| Primary | 1.17 (0.42–3.23) | 1.24 (0.61–2.52) | 0.94 (0.49–1.79) |
| Secondary | 0.94 (0.39–2.31) | 0.97 (0.49–1.94) | 1.11 (0.60–2.04) |
| Tertiary | 1.10 (0.40–3.01) | 0.96 (0.47–1.95) | 0.92 (0.48–1.78) |
| Socioeconomic status |  |  |  |
| Lowest | 1 (Reference) | 1 (Reference) | 1 (Reference) |
| Low | 0.62 (0.31–1.22) | 2.11 (1.27–3.51) | 0.67 (0.44–1.04) |
| Middle | 0.91 (0.48–1.72) | 1.67 (1.00–2.78) | 0.74 (0.47–1.15) |
| High | 0.56 (0.28–1.15) | 2.87 (1.68–4.92) | 0.55 (0.35–0.87) |
| Highest | 0.85 (0.44–1.65) | 2.28 (1.33–3.90) | 0.60 (0.37–0.97) |
| Alcohol intake |  |  |  |
| Never drinker | 1 (Reference) | 1 (Reference) | 1 (Reference) |
| No drinking in last 12 months | 1.21 (0.34–4.29) | 0.30 (0.12–0.76) | 2.00 (1.02–3.94) |
| Drinking in past 12 months | 0.49 (0.26–0.95) | 0.95 (0.63–1.42) | 1.20 (0.84–1.72) |
| Smoking status |  |  |  |
| Never smoked | 1 (Reference) | 1 (Reference) | 1 (Reference) |
| Ever smoked (current/former) | 0.79 (0.37–1.69) | 1.00 (0.65–1.56) | 1.05 (0.69–1.59) |
| HIV status |  |  |  |
| Negative | 1 (Reference) | 1 (Reference) | 1 (Reference) |
| Positive | 1.30 (0.87–1.94) | 1.56 (1.20–2.03) | 0.58 (0.45–0.74) |
| BMI category |  |  |  |
| Underweight | 1 (Reference) | 1 (Reference) | 1 (Reference) |
| Normal | 2.39 (1.04–5.51) | 0.56 (0.30–1.06) | 1.15 (0.72–1.83) |
| Overweight | 2.57 (1.03–6.39) | 0.72 (0.37–1.39) | 0.93 (0.56–1.55) |
| Obese | 3.02 (1.17–7.78) | 0.57 (0.29–1.12) | 1.12 (0.65–1.92) |
| Hypertension |  |  |  |
| No | 1 (Reference) | 1 (Reference) | 1 (Reference) |
| Yes | 0.84 (0.39–1.77) | 0.76 (0.53–1.10) | 1.22 (0.85–1.76) |
| Diabetes |  |  |  |
| No | 1 (Reference) | 1 (Reference) | 1 (Reference) |
| Yes | 1.63 (0.74–3.60) | 0.76 (0.45–1.26) | 1.11 (0.64–1.93) |

HBV: hepatitis B virus; HIV: human immunodeficiency virus; Body Mass Index: BMI; SES - socioeconomic status

Supplementary Table 9. Weighted prevalence of vaccine mediated immunity and infection derived (resolved) immunity based on different anti-HBs thresholds in a rural population in KwaZulu Natal, South Africa.

| **Anti-HBs threshold**  **(mIU/mL)** | **Vaccine-derived immunity,**  **% (95% CI)** | **Infection-derived immunity (resolved infection),**  **% (95% CI)** |
| --- | --- | --- |
| >10 | 8.9 (7.5-10.4) | 31.3 (28.8-33.8) |
| >100 | 2.7 (1.7-3.6) | 19.0 (16.8-21.1) |
| >1000 | <0.1 | <0.1 |

HIV: human immunodeficiency virus; HBV: hepatitis B virus; Anti-HBs**:** HBV surface antibodies; PLWH: person living with HIV; CI: confidence interval

### **Supplementary Figures**

#### Supplementary Figure 1: Distribution of results of HBsAg S/CO >1.0 on Abbott assay.

(A) Distribution of S/CO ratio in all samples reported as HBsAg reactive; 240 samples have S/CO >1.0, of which 193 have S/CO >2.0, 144 have S/CO>5, 129 have S/CO >10 and 101 have SCO>100 (thresholds shown in dashed lines); (B) HBsAg S/CO ratio in 20 samples re-tested on alternative assays with reported status on POCT / Cobas. (C) Samples ranked by HBsAg S/CO ratio showing other markers and final possible interpretation(s) of testing.

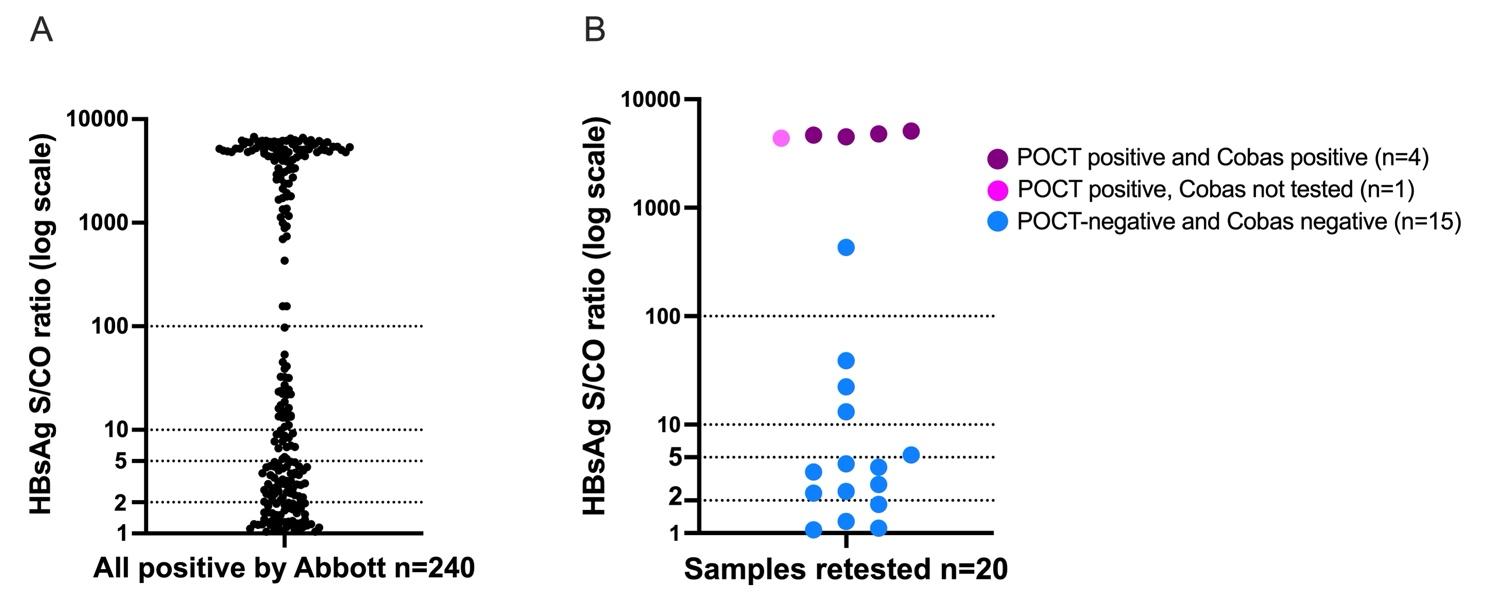

C

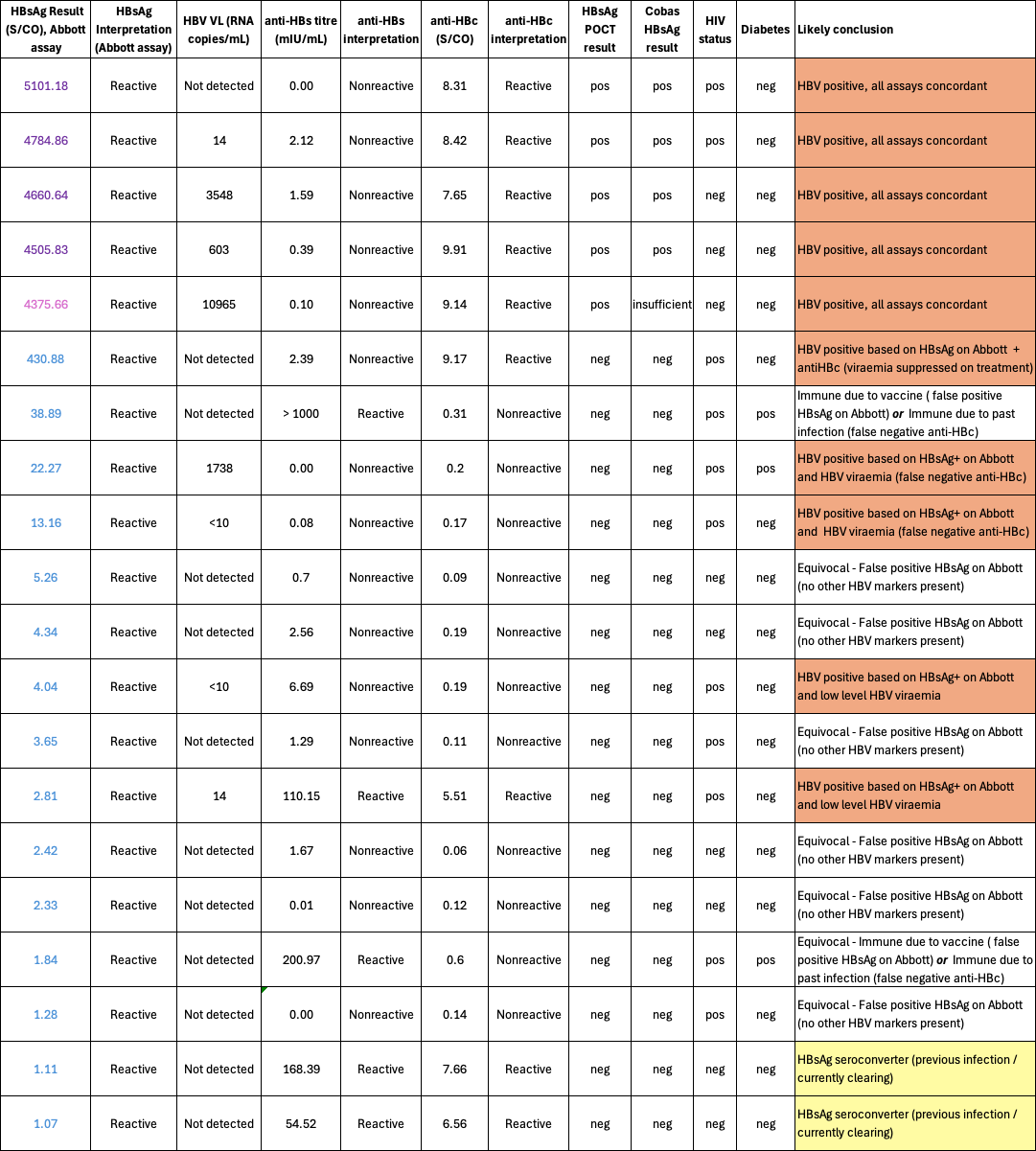

#### Supplementary Figure 2. Distribution of anti-HBs titres in 2200 adults sampled from the Vukuzazi cohort in KwaZulu Natal, South Africa.

Dashed lines indicate thresholds commonly applied in clinical practice to be associated with immunity (anti-HBs >10 potentially immune; 100 likely immune; 1000 robust immunity).[A] Population divided into anti-HBc negative (non-exposed, assumed immunity due to vaccination) and anti-HBc positive (HBV exposed, assumed immunity due to infection). [B] Anti-HBc negative and positive substratified by HIV status. Anti-HBc: HBV core antibodies; anti-HBs: HBV surface antibodies.

[A]

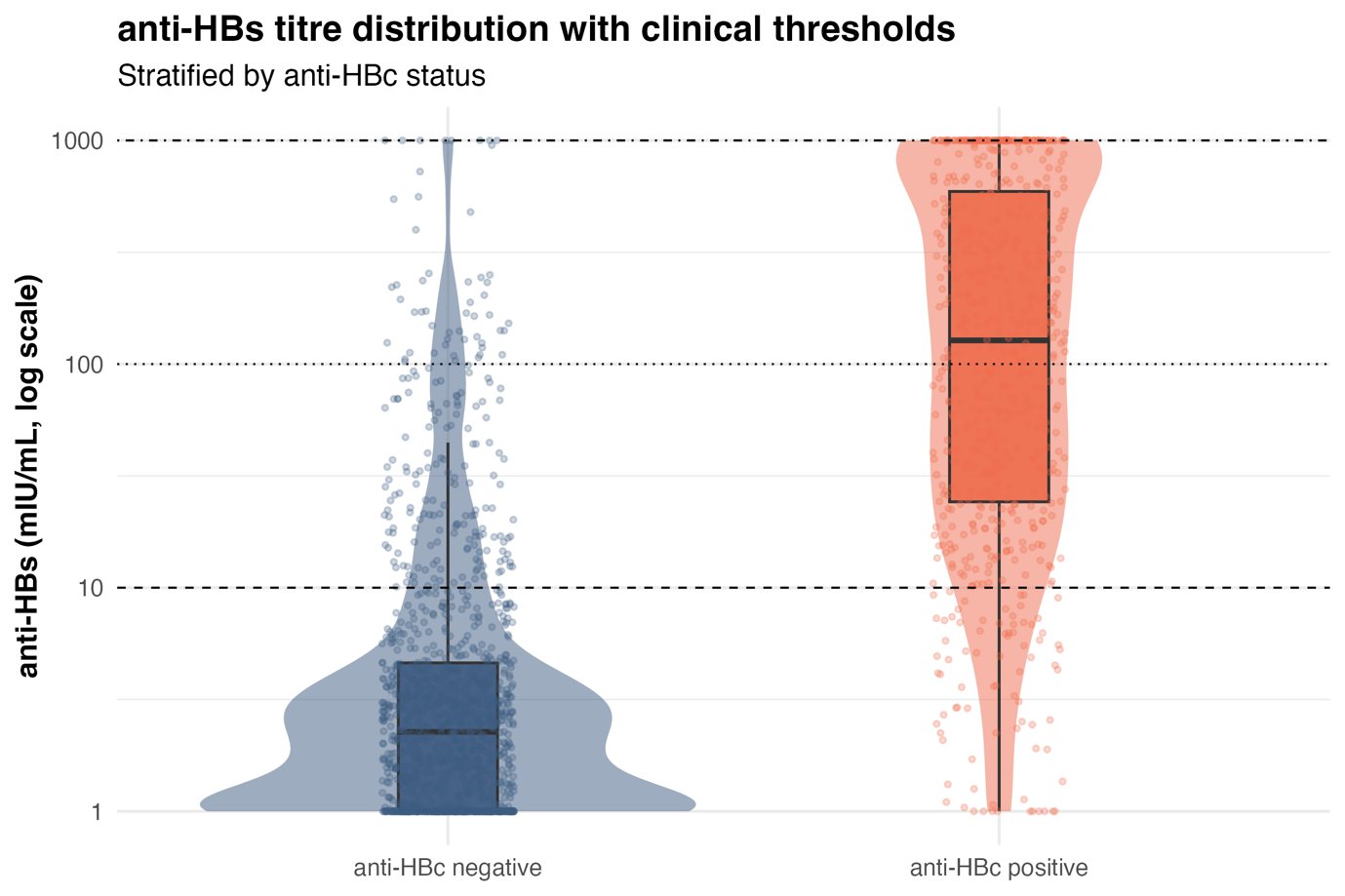

[B]

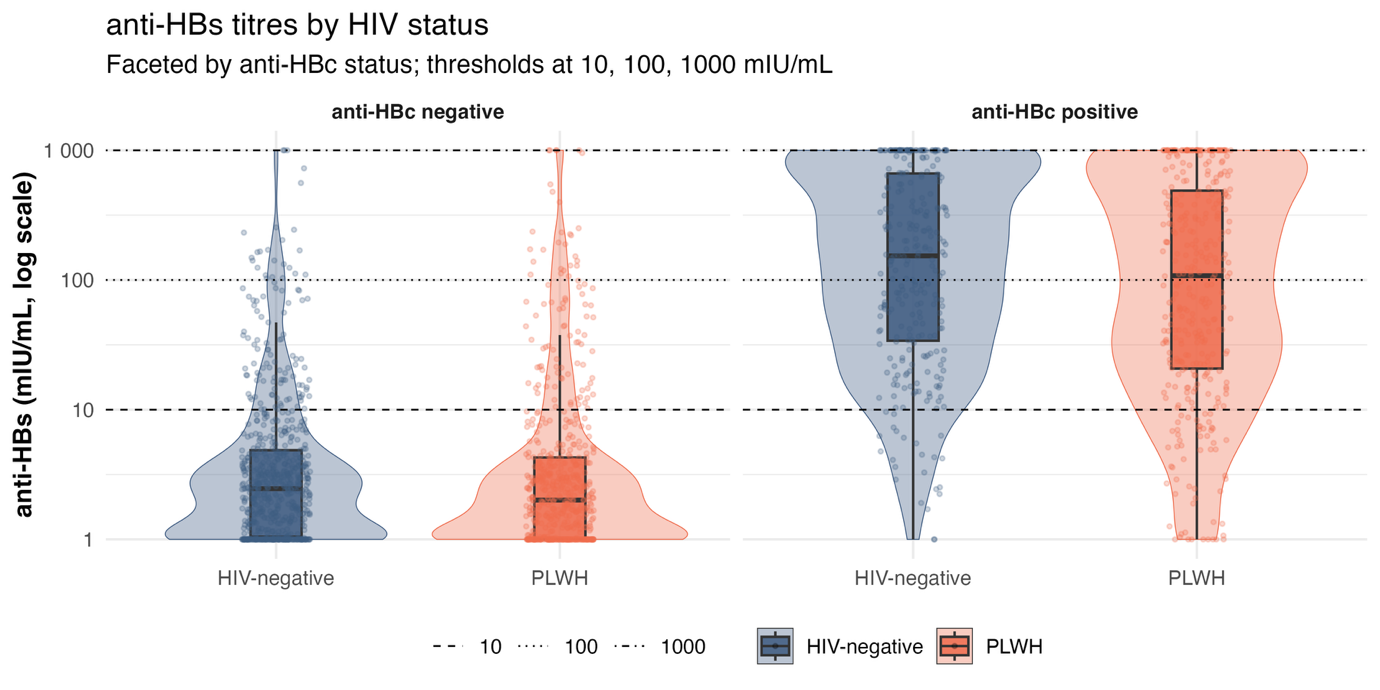

#### Supplementary Figure 3. Forest plot of multivariable analysis of factors associated with HBV serostatus.

Data for 2200 adults sampled from the Vukuzazi cohort in KwaZulu Natal, South Africa, based on Abbott assay with threshold for HBsAg increased to S/CO >10. HBV: Hepatitis B Virus; HIV: human immunodeficiency virus. Odds ratios from analysis adjusted for all variables shown in supplementary table 8.

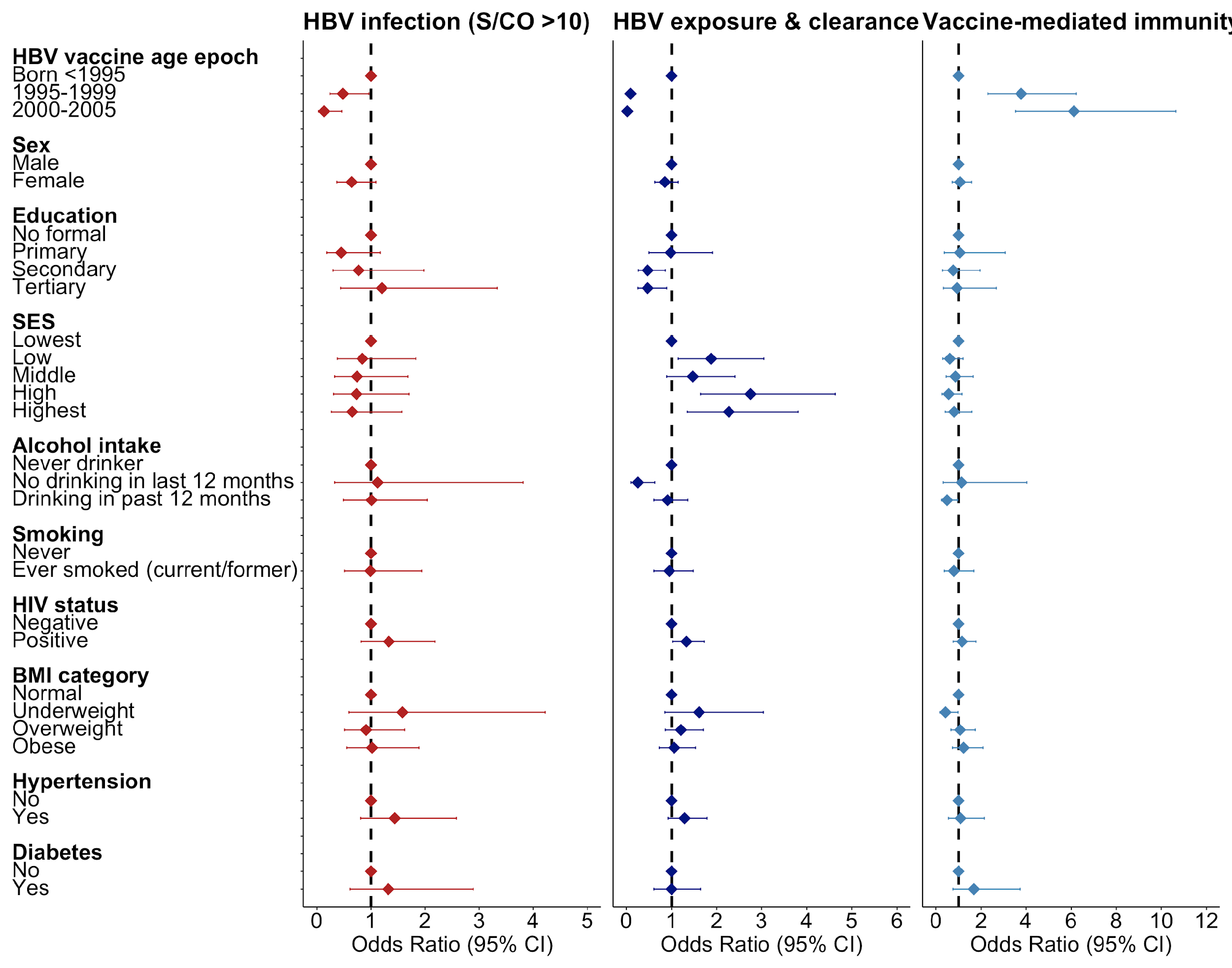
